# Prevalence of SARS-CoV-2 infection in the Luxembourgish population – the CON-VINCE study

**DOI:** 10.1101/2020.05.11.20092916

**Authors:** Chantal J. Snoeck, Michel Vaillant, Tamir Abdelrahman, Venkata P. Satagopam, Jonathan D. Turner, Katy Beaumont, Clarissa P. C. Gomes, Joëlle V. Fritz, Valerie E. Schröder, Anne Kaysen, Lukas Pavelka, Lara Stute, Guilherme Ramos Meyers, Laure Pauly, Maxime Hansen, Claire Pauly, Gloria A. Aguayo, Magali Perquin, Anne-Marie Hanff, Soumyabrata Ghosh, Manon Gantenbein, Laetitia Huiart, Markus Ollert, Rejko Krüger, on behalf of the CON-VINCE study group

## Abstract

**BACKGROUND:** The World Health Organization declared the outbreak of coronavirus disease to be a public health emergency of international concern on January 30, 2020. The first SARS-CoV-2 infection was subsequently detected in Luxembourg on February 29, 2020. Representative population-based data, including asymptomatic individuals for assessing the viral spread and immune response was, however, lacking worldwide.

**METHODS:** Using a panel-based method, we recruited a representative sample of the Luxembourgish population based on age, gender and residency for testing for SARS-CoV-2 infection and antibody status in order to define prevalence irrespective of clinical symptoms. Participants were contacted via email to fill an online questionnaire before biosampling at local laboratories. Participants provided information related to clinical symptoms, epidemiology, socioeconomic and psychological assessments and underwent biosampling, rRT-PCR testing and serology for SARS-CoV-2.

**RESULTS:** A total of 1862 individuals were included for our representative sample of the general Luxembourgish population. We detected an ongoing SARS-CoV-2 infection based on rRT-PCR in 5 participants. h Four of the SARS-CoV-2 infected participants were oligosymptomatic and one was asymptomatic. Overall, 35 participants (1.97%) had developed a positive IgG response, of whom 11 self-reported to have previously received a positive rRT-PCR diagnosis of SARS-CoV-2 infection. Our data indicate a prevalence of 0.3% for active SARS-CoV-2 infection in the Luxembourgish population between 18 and 79 years of age.

**CONCLUSIONS:** Luxembourgish residents show a low rate of acute infections after 7 weeks of confinement and present with an antibody profile indicative of a more recent immune response to SARS-CoV-2. All infected individuals were oligo- or asymptomatic. Bi-weekly follow-up visits over the next 2 months will inform about the viral spread by oligo- and asymptomatic carriers and the individual changes in the immune profile.

## Introduction

On March 11, 2020, the World Health Organization (WHO) declared the ongoing outbreak of coronavirus disease (COVID-19) a pandemic (World Health Organization (WHO), 2020b). The pathogen responsible for COVID-19, severe acute respiratory syndrome coronavirus 2 (SARS-CoV-2), was first described in Wuhan, Hubei Province, China (Huang et al., 2020). By May 6, 2020, the illness had rapidly spread to over 210 countries, affecting over 3 million people and claiming more than 200,000 lives worldwide (World Health Organization (WHO), 2020a).

The first SARS-CoV-2 infection in Luxembourg was detected on February 29, in a person who had just returned from Italy. Luxembourg is a country with 626,000 inhabitants in the heart of Europe and has borders with Belgium, France, and Germany (Statec; 2020). It is highly connected to its neighbors, with more than 200,000 cross-border employees commuting to Luxembourg. On March 12, the government communicated measures taken to tackle the spread of the Coronavirus. These measures entered into force on March 16. As in many other countries, all educational institutions were closed, employees were encouraged to work from home whenever possible, all non-essential activities were cancelled or closed, including all commercial and business activities involving direct contact with customers (except essential products and services). Travel was restricted for the general public (with limited exceptions such as helping and caring for elderly or disabled people, or travelling to work, and border controls were implemented). By May 6, 50533 individuals had been tested, with 3851 individuals tested positive for SARS-CoV-2 and the disease accounted for 98 deaths in Luxembourg so far (Government of the Grand Duchy of Luxembourg, 2020: https://msan.gouvernement.lu/en/dossiers/2020/corona-virus.html).

In addition to the direct effects of the pandemic, the confinement and quarantine measures linked to the coronavirus pandemic also have a substantial socio-economic and psychological impact. Besides initiating a global recession, psychological counselling services opened worldwide and will soon demonstrate the psychological cost of the pandemic. Further analyses over time will provide insights into the expansion of the socio-economic and psychological impact of the coronavirus pandemic.

First studies currently aim at better understanding the dynamics of the pandemic and to improve measures to confine the spread of SARS-CoV-2 in the absence of effective medication or of vaccines (Gudbjartsson et al., 2020). Here the role of asymptomatic SARS-CoV-2-positive individuals for the propagation becomes more and more important (Streeck et al., 2020; Lai et al., 2020; Bai et al., 2020). Systematic screening of a representative population irrespective of clinical symptoms may detect asymptomatic persons who are expected to play an important role in the disease transmission and inform about the overall infection rate in a population. We used reverse-transcription polymerase chain reaction (rRT-PCR) to identify asymptomatic or oligosymptomatic carriers of SARS-CoV-2 and serology-based testing to identify participants that had developed an immune response. In addition, questionnaires on clinical symptoms, epidemiological, and socioeconomic factors were used to better understand the nature, dynamics of spread, and transmission along with the prevalence of the virus in the population.

## Materials and Methods

### Study design

This study is a nation-wide, observational study aiming to define the prevalence of SARS-CoV-2 infections in the Luxembourgish population. Here we present the baseline assessment of the CON-VINCE cohort. To capture the dynamics and impact of the virus spreading over time, the cohort will be follow-up over 12 months. An intensive data and sample collection will be performed every 2 weeks for the first 2 months (5 times in total) with a final follow-up 1 year after the participants’ inclusion in the study. At each collection time-point, blood, nasal and oropharyngeal swabs are collected, and participants also fill in questionnaires on epidemiological and clinical data as well as socioeconomic and psychological well-being. The provision of a stool sample at each biosampling time point is optional for all participants. To allow for data harmonization and international collaboration the overall data set includes questions recommended by the International Severe Acute Respiratory and emerging Infection Consortium (ISARIC), hosted by the Centre for Tropical Medicine and Global Health at the University of Oxford (WHO), and the Weizmann Institute (Rossman et al., 2020) adapted to the Luxembourgish environment (Börsch-Supan et al., 2013; Börsch-Supran, 2019).

The study design (**Figure 1**) accounts for the need to recruit a representative sample of the Luxembourgish population (>18 years old) within a short time frame in the context of the already existing confinement measures. All participants were tested for SARS-CoV-2 by rRT-PCR. Additionally, serological testing for virus-specific antibodies (IgA and IgG against SARS-CoV-2) was performed. Participants who were either asymptomatic or oligosymptomatic, from the clinical point of view, are being followed up longitudinally regardless of the SARS-CoV-2 positive or negative status. Symptomatic individuals positive for SARS-CoV-2 with COVID-19 disease contribute to the baseline assessment but are not followed-up.

**Figure 1:**
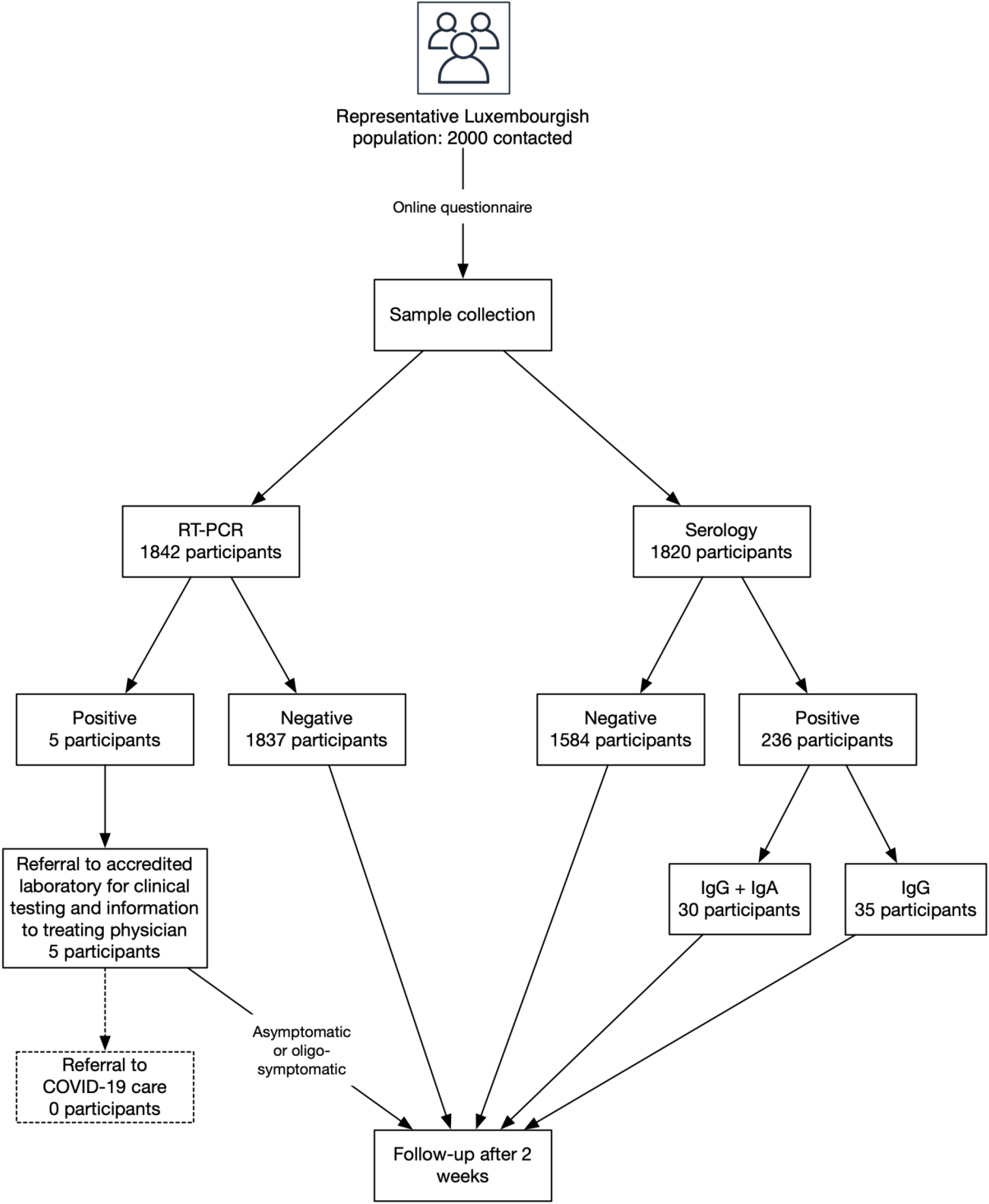
Study design and testing. A representative sample of the Luxembourgish population was invited to join the study. Following the completion of an online questionnaire and biosampling, rRT-PCR and serology were performed. Asymptomatic and oligosymptomatic participants (either positive or negative for the virus) were followed up on a bi-weekly basis for a total of 4 times. A final follow-up will take place after one year.

The baseline questionnaire captured demographic data, medical history and behavioral and psychological data. Demographic data included age, gender, origin, residential areas as well as marital status, number of children, the household composition, and the age of the household members. As socio-economic-status is intimately linked to the incidence and severity of respiratory tract infections, we obtained information on educational level, professional background, current employment status, income, and the house-ownership.

Subjects provided information on their medical history, including cardiac, hepatic, metabolic, pulmonary, neurological, hematological and oncological comorbidities, and gynecological history. Prior history of allergies and smoking were recorded. A self-reported description of chronic medication taken regularly was mandatory. COVID-19-related data included whether the participants have already been tested for the SARS-CoV-2, indicating the date and the result of the test. Additionally, we invited subjects to document if they had travelled to an area with confirmed SARS-CoV-2 infections during the 14 days prior to their participation in the study. Environmental conditions of the household were obtained (e.g., the possibility of quarantine of one household member, etc.).

To assess psychological and behavioural factors or changes during the pandemic, we asked participants to quantify physical activity, the frequency of leaving the house, alcohol consumption, screen time, as well as social contact through technological devices. Their compliance with the recommendations and restrictions issued by the Luxembourgish government during the pandemic was measured by a series of questions compiled from the WHO and Luxembourg Health Directorate guidelines (WHO 2014, Luxembourg Health Directorate, 2020). Depressive symptoms are and further will be assessed in the multiple follow-up questionnaires using the Center for Epidemiologic Studies Depression Scale (CES-D Scale) (Radloff, 1977). Presence and severity of anxiety is measured in the multiple follow-up questionnaires using the seven items Generalized Anxiety Disorder Assessment (GAD-7) (Spitzer et al., 2006). Participants’ perception of their psychosocial stress level was assessed using the 4-item Perceived Stress Scale (PSS-4) on a 5-point Likert scale (Cohen et al., 1983). Social isolation and loneliness were assessed with the 3-item UCLA Loneliness Scale (short version) (Hughes et al., 2004). The Brief Resilience Scale (BRS) (Smith et al., 2008) measures resilience or the ability to recover from a stressful period or event using 6 items, scored on a 5-point Likert scale. Five personality traits (Extraversion, Agreeableness, Conscientiousness, Emotional Stability and Openness) are assessed using the Big Five Inventory-10 (BFI-10) (Rammstedt and John, 2007).

The bi-weekly follow-up questionnaires will cover participants’ present health and psychological status. Current health status will focus on signs and symptoms associated with SARS-CoV-2 during the intervening period and medication (e.g., paracetamol, cough medicine, NSAIDs) for symptom relief. Psychological data that will be collected during the follow-up questionnaires include CES-D, GAD-7, PSS-4 and UCLA loneliness scale. BRS and BFI-10 are administered again during the yearly follow-up. As social adversity has been linked to respiratory tract infections (Elwenspoek et al., 2017) the final questionnaire includes the 28-item Childhood Trauma Questionnaire (Bernstein et al., 2003), to retrospectively assess distress during childhood together with a questionnaire covering the principal psychosocial stressors in adulthood such as divorce, job loss or the death of a family member (Turner at al., 2020). The questionnaires, as well as the questionnaire schedule, are included in **Supplementary Material 1**.

The study was approved by the national research ethics committee (Comité National d’Ethique de Recherche, CNER), under reference 202004/01, and by the Luxembourgish Ministry of Health under reference 831×6ce0d. The study has been submitted for registration on ClinicalTrials.gov (NCT04379297).

### Population screening

The CON-VINCE study was launched on April 15, and participants randomly selected from a representative panel, which consented to the study and fulfilled the inclusion criteria, were enrolled from April 15 until May 5, 2020. Participants filled the online questionnaire and accomplished biosampling in approved diagnostic laboratories spread all over the country. Participants vulnerable to SARS-CoV-2 infection were sampled at home. A total of 1862 participants successfully completed the questionnaire and were biosampled.

### Sample size calculation

Given the rapidly evolving incidence of infected cases on April 10, when the study protocol of CON-VINCE was established and based on an unknown number of asymptomatic people, a prevalence of 50% of cases was assumed, that would lead to the largest sample size. It would allow estimating any other prevalence figure.

Assuming a 95% confidence interval and a precision of 2.5% around the estimate of prevalence, the required sample size was a minimum of 1537 participants. The chosen sampling strategy for the sample to be representative of the general population was to stratify by gender, age categories (10 years from 18 years and above), and electoral districts. To compensate for non-response and potential drop-outs during the study, over 2000 individuals were invited to join the study.

### Sampling

According to the National Institute of Statistics and Economic Studies of the Grand Duchy of Luxembourg (Statec), the national population aged 18 years and over was 514,921 at the beginning of the study. This constituted the sampling frame.

An equal allocation probability was used (chance for all individuals of the same age category and gender to be selected) proportional to size (of the population) without replacement (the same individual could not be selected twice). The selection probability for unit i (for example 60-69y) in stratum h (for example men) equaled nhZhi, where nh was the sample size for stratum h, and Zhi was the relative size of unit i in stratum h. The relative size equaled Mhi/Mh, which was the ratio of the size measure for unit i in stratum h (Mhi = number of men aged 65-69y) to the total of all size measures for stratum h (Mh = number of men) (SAS Institute Inc, 2009).

Due to constraints related to the emerging pandemic, the sample of participants was enrolled through the use of a non-probabilistic web panel (unknown probability to opt-in) of 18,000 members by a survey company to access participants within the sampling plan. A deterministic random bit generator (DRBG) within strata was used to apply the equal allocation probability proportional to size.

### Acquisition and Preparation of Samples

The first round of biosampling was performed from April 16 onwards at routine medical diagnostic laboratories (BioneXt, Ketterthill and Laboratoires Réunis) throughout Luxembourg using standardised study collection kits containing all the required materials provided by the Integrated Biobank of Luxembourg (IBBL). Collection kits included the mandatory blood samples and nose and throat swabs, as well as an optional stool collection kit (**Supplementary Table 1**). Following collection, blood, nose and throat swabs were maintained at 2-8°C and transported within 24h to the IBBL for processing. Participants collected stool samples at home and sent them in provided transport boxes to the IBBL using the regular postal service. Blood and stool samples were processed in the IBBL. Swab samples were transferred to the Laboratoire National de la Santé (LNS) for rRT-PCR SARS-CoV-2 testing (**Supplementary Table 1**). Serum was transferred at −20°C to the Department of Infection and Immunity of the Luxembourg Institute of Health (LIH) for serology testing. Leftover swab samples and residual RNA from swab samples were returned to IBBL for aliquoting and storage, within 24h following the initial transfer to LNS. The sampling procedure will be maintained for the follow-up visits.

### Data Integration, Storage and Access

As part of our Case Report Forms (CRFs) (**Supplementary Material 1**) was based on ISARIC COVID19 Core CRF (https://isaric.tghn.org/COVID-19-CRF/), our data is aligned with international standards and can be easily harmonized and pooled with other studies facilitating cross-study analysis. Encrypted pseudonymised raw data has been deposited in the Luxembourg Centre for Systems Biomedicine (LCSB) secure repository. Upon arrival, all pseudonymised questionnaire and clinical data is curated and mapped to associated rRT-PCR and serology data through each participant’s pseudonymised ID with corresponding sample kit ID (subject-sample-mapping) and maintained in REDCap (www.project-redcap.org), personal information was maintained separately in a proprietary secured server (SMASCH). REDCap provides a complete audit trail for study data (https://www.project-redcap.org). This integrated, curated and processed data is available through an access-controlled study-specific data integration and analysis platform (ADA). The ELIXIR-Luxembourg node (ELIXIR-LU http://elixir-luxembourg.org) will provide long-term hosting, ensuring the sustainability of CON-VINCE data. Data flows are highlighted in **Figure 2**).

**Figure 2:**
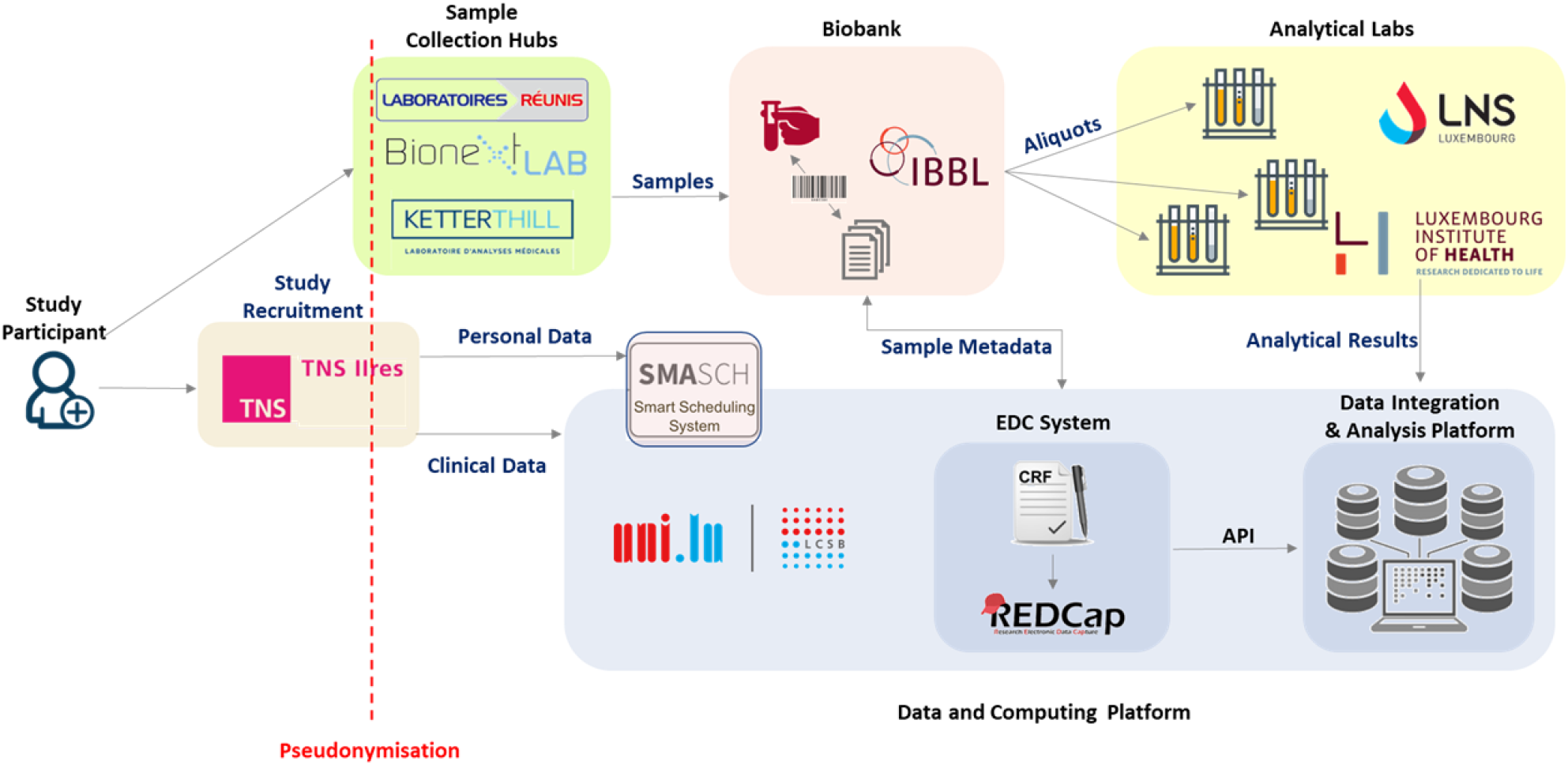
Data and sample flow in CON-VINCE cohort. Study participants are recruited by a survey company (TNS-Ilres) based on a large representative panel of residents of Luxembourg. Participant personal data is securely collected and can only be accessed by the clinical study team. Each participant’s personal record is assigned with a pseudonym. All data is collected by a protected web-based interface and transferred along with the pseudonym to the Data and Computing Platform hosted at the LCSB in a secure data center. Biosamples are collected at different sample collection hubs across Luxembourg and shipped to IBBL for sample processing and biobanking. Biosamples are analysed at the Laboratoire National de Santé (LNS) and the Luxembourg Institute of Health (LIH) and the analysis results together with biosample annotations and barcodes are recorded in REDCap. Within the Data and Computing Platform, the pseudonymized data and results from biological analyses from REDCap is accessed by the Data Integration and Analysis Platform (Ada) via an Application Programming Interface and integrated results with clinical data made available through its secure and access-controlled web-application.

### RT-PCR

Automated RNA extraction was performed using a STARMag 96 × 4 Universal Cartridge Kit (Seegene) for all swabs. SARS-CoV-2 detection was carried out using the Allplex 2019 n-CoV Assay (Seegene) according to the manufacturer’s instructions. The Allplex 2019 n-CoV assay amplifies specific regions in the RdRP, N genes (specific SARS-CoV-2 detection) and E gene (pan-*Sarbecovirus* detection). Inconclusive results, i.e. samples where only the RdRP or the N gene was amplified, a second manual RNA extraction using the QIAamp viral RNA minikit (Qiagen, Venlo, the Netherlands) followed by duplicate RNA testing with in house assays including the E gene rRT-PCR (Corman et al., 2020), N gene (N1 target) rRT-PCR and human RNAse P rRT-PCR as sample quality control (CDC), using TaqPath 1-Step RT-qPCR Master Mix (Life Technologies, Merelbeke, Belgium). Viral RNA from BetaCoV/Germany/BavPat1/2020 strain (Ref 026N-03889), kindly provided by the Charité-Universitätsmedizin Berlin (Corman et al., 2020) through the European Virus Archive Global platform, was used as positive control. Inconclusive samples were also screened with the FTD SARS-CoV-2 assay (Fast Track Diagnostics, Esch-sur-Alzette, Luxembourg), which detects N and ORF1ab genes of SARS-CoV-2. As outlined in **Figure 3**, all samples that were only positive for N gene in the original rRT-PCR, were considered positive when a Ct value < 40 was obtained for at least one replicate for 2 viral genes in 2 different second-round rRT-PCRs on the same sample.

**Figure 3:**
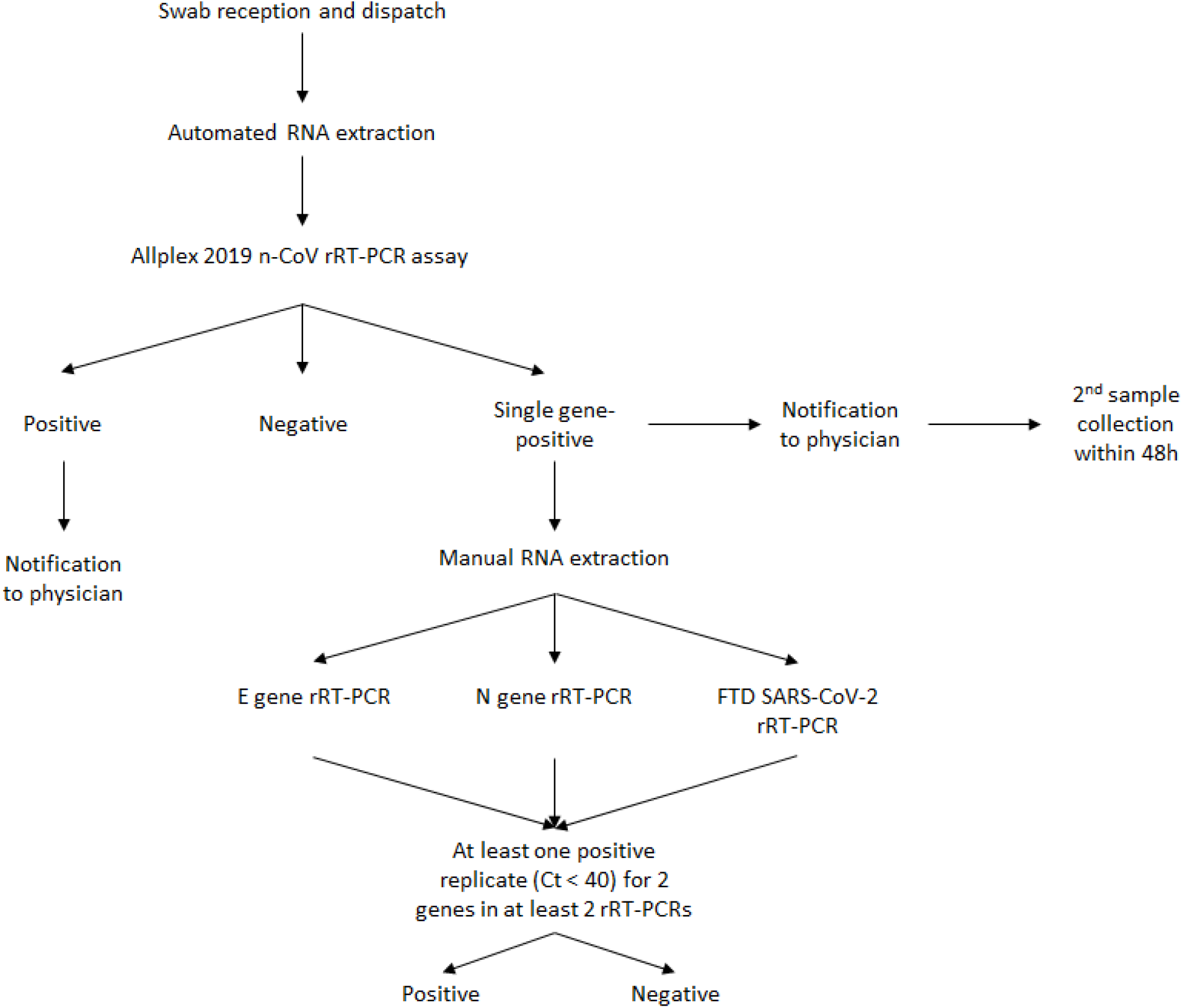
Algorithm for swab testing by rRT-PCR and decision tree

### Serology

Anti-SARS-CoV-2 IgA and IgG were determined by CE-labelled enzyme-linked immunosorbent assay (ELISA) kits (most recent versions of Euroimmun Anti-SARS-CoV-2 ELISA IgA and Euroimmun Anti-SARS-CoV-2 ELISA IgG) according to the manufacturer’s instructions and as described by others (Streeck et al., 2020). The optical density (OD) was measured at 450 nm from which background OD measured at 650 nm was deducted. OD ratios were calculated by dividing the resulting OD by the OD of the calibrator, which is included in the kit. In house quality controls, prepared to give an expected OD ratio of approximately three times the threshold for positivity, were included in all assays. As per the kit recommendations, samples with OD ratios <0.8 were considered negative, OD ratios ≥1.1 were considered positive and samples with intermediate OD ratios (≥0.8, <1.1) were judged borderline positive.

A cohort of anonymised archived sera collected from adults during the two winter seasons (cohort A, 2018, n=92; 2019, n=93) prior to the start of the COVID-19 pandemic were tested by both IgA and IgG ELISAs to assess their specificity. All sera had been collected for other purposes than respiratory disease diagnosis. A second cohort of anonymized sera (cohort B, 2020, n=37) was collected at the Centre Hospitalier de Luxembourg (CHL) from patients with documented COVID-19 disease and positive rRT-PCR results. The delay between onset of symptoms and blood drawing ranged from 1 to 26 days. Those sera contributed to evaluating the ELISA sensitivity. All sera with intermediate results were considered positive when calculating specificity and sensitivity.

### Statistical analyses and data presentation

Prevalence of the SARS-CoV-2 RNA infection was measured by rRT-PCR. IgA and IgG serology was also translated into seroprevalence. The infection rate was evaluated through previously reported and current rRT-PCR and IgG positivity. Prevalence estimates were calculated by accounting for the design of the study, namely weights from the sampling strategy as defined by the stratification variables (gender, 10 years age categories and electoral district).

A representativeness evaluation was carried out to verify that the proportions estimated from the sample can be extrapolated to the general population. 95% confidence intervals (95%CI) were given. Sampling proportions were calculated for each strata of the sample of participants enrolled in the study as well as in the general population. The ratio of these proportions provided the weights for post stratification.

In the sense of sensitivity analyses, sampling weights were also calculated for “cantons” which is a more granular strata than electoral district.

The formula for infection rate was as follows: past or current positive PCR or IgG positive or intermediate divided by the total sample population (N=1835).

Prevalences were calculated with SAS v9.4 (SAS institute, Cary, NC, USA) except the IR and IFR where R was used with the svycipro function (Survey package) that calculates 95% confidence intervals for proportions with logit » method. Figures were prepared in R studio (R Core Team, 2019) using the tidyverse (Wickham et al. 2019), ggbeeswarm (Clarke and Sherril-Mix, 2017) and ggpubr (Kassambara, 2020) packages.

## Results

### Cohort description

The recruitment of Luxembourg residents for the CON-VINCE study started on April 15 and was concluded on May 5 after the inclusion of 1862 individuals, thereby exceeding the minimum of 1537 participants calculated for assessing prevalence. All individuals that accomplished the baseline questionnaire online underwent biosampling. Viral rRT-PCR was performed in 1842 participants and SARS-CoV-2 specific serology for IgA and IgG was performed in 1820 individuals.

The basic epidemiological features of the study participants, such as gender, age, education, number of persons sharing the same household, and residency within the Luxembourg territory are described in **Table 1**.

**Table 1:**
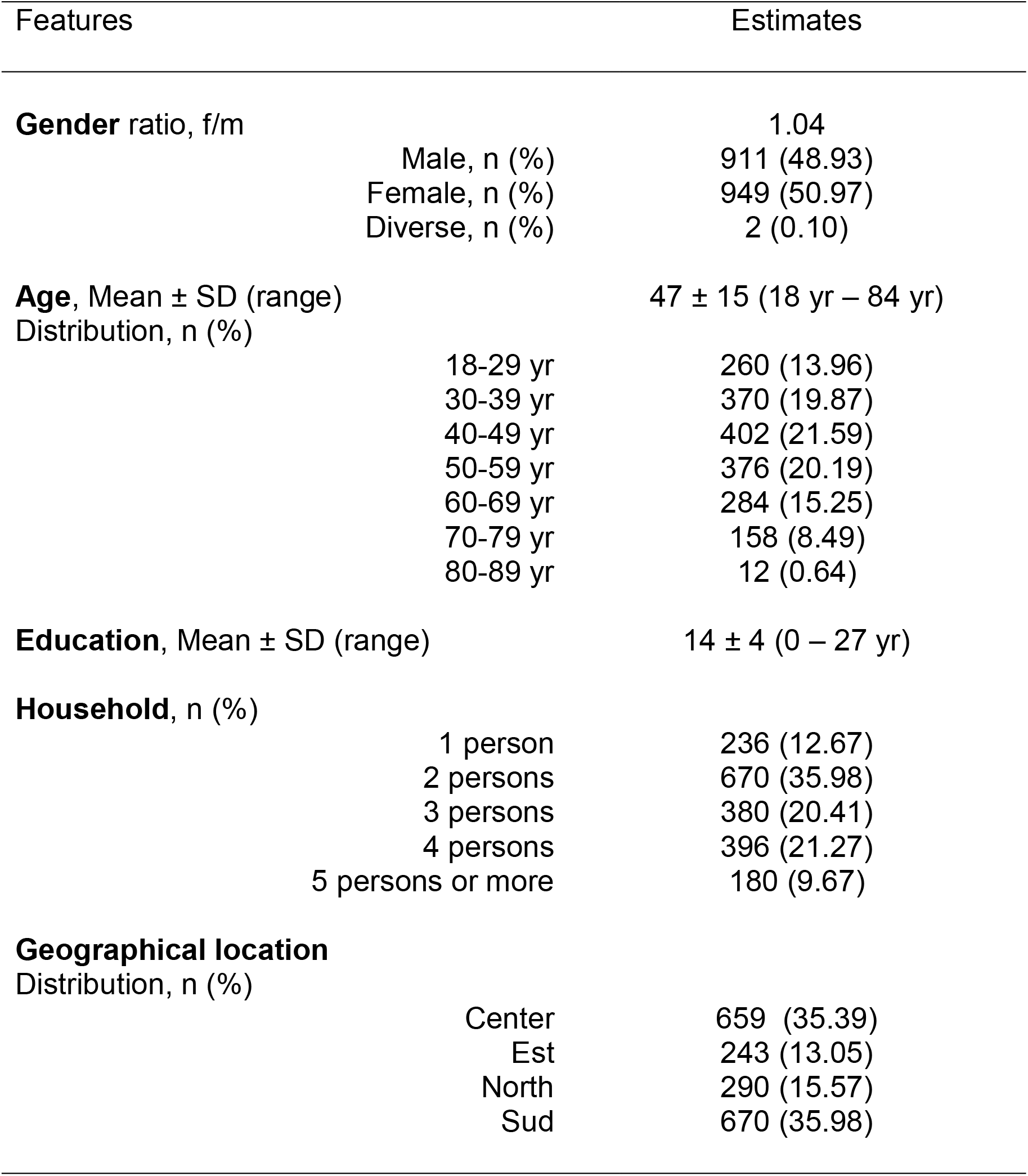
Population features of the 1862 participants

### Sensitivity and specificity of SARS-CoV-2 IgA and IgG ELISAs

Two serum cohorts were used to assess the specificity and sensitivity of both IgA and IgG ELISAs. In specificity cohort A, 20/185 (10.8%) and 4/185 (2.2%) sera reacted in IgA and IgG ELISAs (positive and intermediate; **Figure 4**), leading to a specificity of 89.2% and 97.8% respectively. Only one serum was IgA and IgG positive, providing an increased specificity of 99.5% when combining both results.

**Figure 4:**
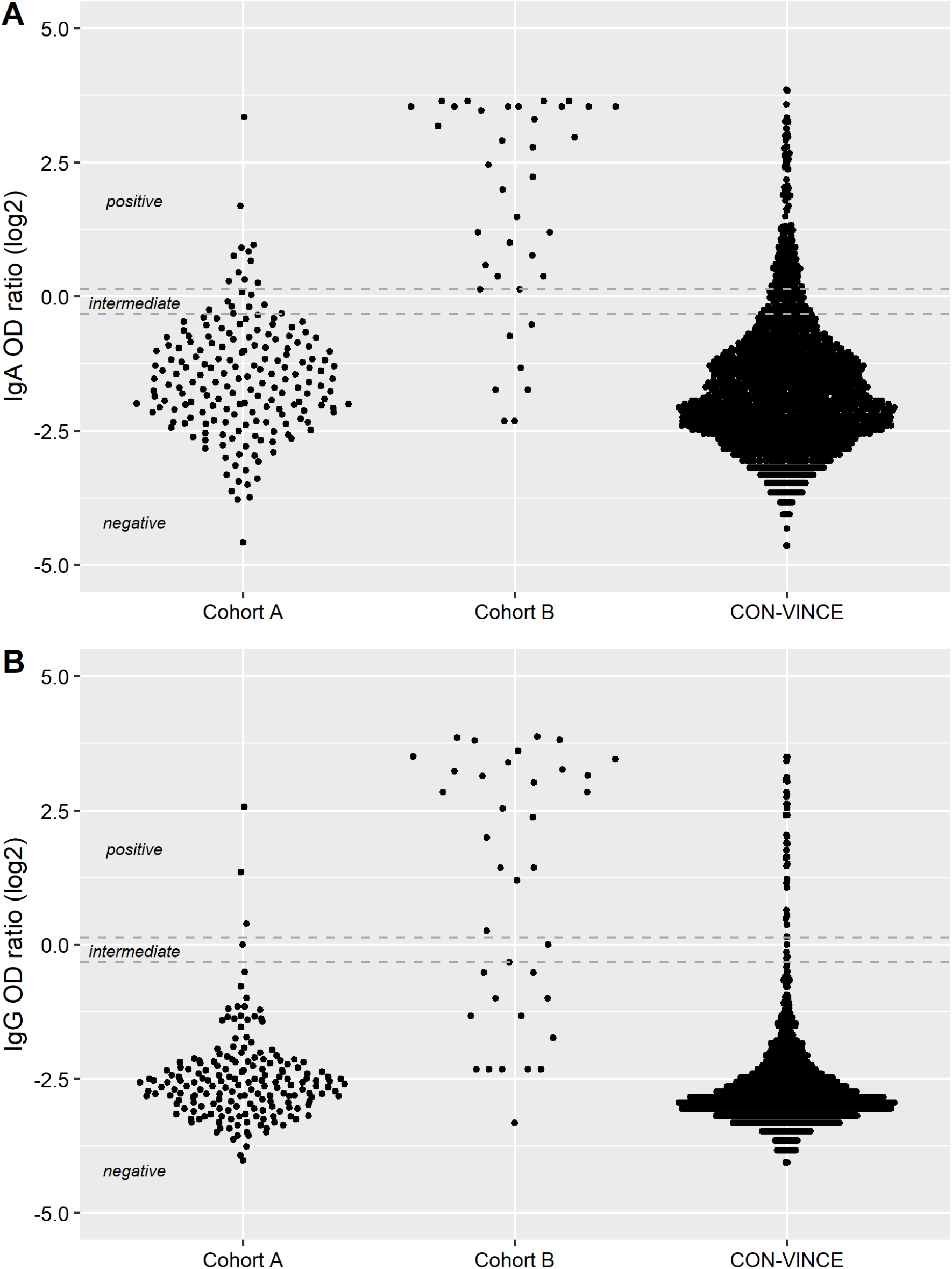
Density plots of IgG (panel A) and IgA (panel B) OD ratios (log2 transformation) of patient sera collected in 2018-2019 (specificity cohort A, n=187), COVID-19 convalescent patients (sensitivity cohort B, n=37) and CON-VINCE participants (n= 1820). When measuring IgA antibodies, the maximum OD read-out was used for 11/37 COVID-19 convalescent patients for which saturation was reached (panel B, cohort B, Log2 OD ratios > 3.5). Dotted lines represent thresholds for categorizing samples into negative, intermediate and positive.

Cohort B consisting of hospitalized COVID-19 patients was used to estimate assay sensitivity in relation to the time delay after symptom onset (**Table 2**). IgA and IgG ELISA sensitivity reached 92.9% and 85.7% at 15 days (d15) after symptom onset, while the combination of IgA and IgG results provided a sensitivity of 85.7% (**Table 2**).

**Table 2:** Sensitivity of Euroimmun IgA and IgG ELISAs and combined IgA and IgG interpretation

| Delay after symptom onset | No. of pos*/total No. of samples in this category (sensitivity in %) |  |  |  |
| --- | --- | --- | --- | --- |
|  | IgA | IgG | IgA and IgG | IgA and/or IgG |
| > 5 days | 27/33 (81.8) | 23/33 (69.7) | 21/33 (63.6) | 29/33 (87.9) |
| > 10 days | 22/25 (88.0) | 21/25 (84.0) | 19/25 (76.0) | 24/25 (96.0) |
| > 15 days | 13/14 (92.9) | 12/14 (85.7) | 12/14 (85.7) | 13/14 (92.9) |

\* Sera with intermediate results were considered positive

### Serological screening at baseline

The presence of anti-SARS-CoV-2 IgA and IgG was assessed in all sera using the CE-labeled Euroimmun assays. Overall, 201/1820 (11.0%), and 35/1820 (1.9%) participants had IgA or IgG antibody levels above the threshold considered for positivity in our study (OD ratio ≥0.8), respectively. Among those, 30 (1.6%) participants were positive for both IgA and IgG (**Table 3**; **Figure 5**). A positive correlation between IgA and IgG OD ratios was observed (all data, Spearman correlation coefficient *r=0.362, p<0.001; IgA positive and IgG positive only, r=0.855, p<0.001*).

**Figure 5:**
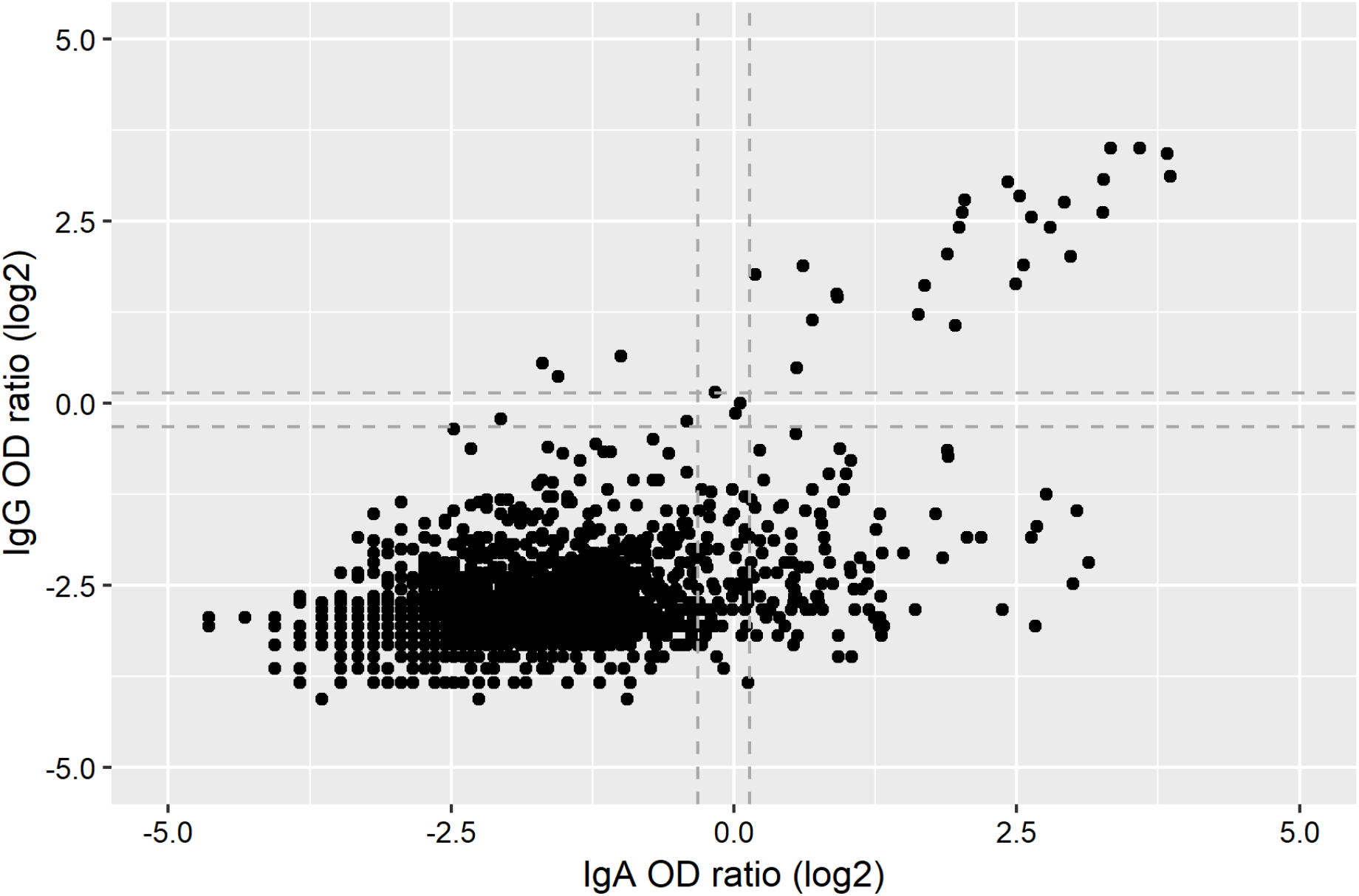
Distribution of IgA and IgG OD ratios (log2 transformation) from 1820 CON-VINCE participants. Dotted lines represent thresholds for categorizing samples into negative, intermediate and positive.

**Table 3:** Proportions of participants with detectable anti-SARS-CoV-2 IgA and IgG antibody response

| IgG status | No. of sera with a given IgA status according to the IgG status (%) |  |  |
| --- | --- | --- | --- |
|  | IgA negative | IgA positive | Total |
| IgG negative | 1614 (88.7) | 171 (9.4) | 1785 (98.1) |
| IgG positive | 5 (0.3) | 30 (1.6) | 35 (1.9) |
| Total | 1619 (89.0) | 201 (11.0) | 1820 (100.0) |

### rRT-PCR screening at study baseline

A total of 1842 upper respiratory tract swabs were analyzed by Allplex 2019 n-CoV Assay. Six (0.3%) gave inconclusive results, i.e. a positive result was only detected by N gene rRT-PCR. Low levels of SARS-CoV-2 viral RNA, as shown by the high Ct values (**Table 4**), were confirmed for 5/6 participants after retesting by additional rRT-PCRs (**Figure 3**). Despite the very low viral RNA concentrations, the repeated viral RNA detection combined with the serological response observed strongly suggested that these five participants had been infected with SARS-CoV-2. Apparently, they were in the phase of clearing the viral infection and mounting an antibody response. This is further evidenced for participant 3, who already tested positive 25 days prior to study enrolment. All participants were oligo- or asymptomatic, 2 participants had contact with a COVID-19 household member while one participant had traveled in the last 14 days (**Table 6**).

**Table 4:** Overview of combined rRT-PCR results and antibody response for six participants with initial single gene-positive rRT-PCR outcome

| Participant (P) | Ct values of first detection rRT-PCRs <sup>a</sup> | No. of replicates detected by confirmatory rRT-PCRs (Mean Ct value) <sup>b</sup> |  |  | RT-PCR consensus result at v0 | RT-PCR results 48h after v0 | Serology results |  |
| --- | --- | --- | --- | --- | --- | --- | --- | --- |
|  |  | In house E gene | In house N gene | FTD |  |  | IgA OD ratio (interpretation) | IgG OD ratio (interpretation) |
| P1 | 36.00 | 1/2 (37.36) | 2/2 (36.09) | 2/2 (36.89) | Positive | n.a. | 1.0 (Intermediate) | 0.9 (Intermediate) |
| P2 | 35.10 | 1/2 (36.16) | 2/2 (35.42) | 2/2 (36.65) | Positive | Negative | 4.0 (Positive) | 5.3 (Positive) |
| P3 | 35.80 | 1/2 (35.33) | 2/2 (36.59) | 1/2 (39.51) | Positive | Negative | 10.1 (Positive) | 11.3 (Positive) |
| P4 | 34.15 | 2/2 (35.12) | 2/2 (34.88) | 2/2 (35.18) | Positive | n.a. | 5.8 (Positive) | 7.2 (Positive) |
| P5 | 35.77 | 0/2 | 0/2 | 0/2 | Currently negative | n.a. | 0.4 (Negative) | 0.2 (Negative) |
| P6 | 36.6 | 1/2 (36.63) | 2/2 (35.30) | 2/2 (37.30) | Positive | n.a. | 5.4 (Positive) | 8.2 (Positive) |

<sup>a</sup> All inconclusive samples were negative in Allplex 2019-nCoV Assay E gene and RdRp gene targets, but positive in N gene.
<sup>b</sup> Confirmatory testing was done by in house generic rRT-PCR assays for the E gene and N gene targets and in addition by the FTD SARS-CoV-2 assay using N gene and ORF1ab targets.

**Table 5:** Overview of self-reported symptoms and self-reported possible source of SARS-CoV-2 exposure in the last 14 days prior to enrolment for six study participants with initial single gene-positive rRT-PCR outcome

| Participant | Self-reported symptoms | Self-reported possible source of exposure to SARS-CoV-2 |
| --- | --- | --- |
| P1 | fatigue or malaise, headache | none |
| P2 | cough, runny nose, pink eye | travel history documented COVID-19 cases |
| P3 | cough | contact with COVID-19 household member |
| P4 | fatigue or malaise, headache | none |
| P5 | none | none |
| P6 | none | contact with COVID-19 household member |

### rRT-PCR screening prior to enrolment

A total of 138/1862 (7.4%) participants had been tested for SARS-CoV-2 by rRT-PCR prior to study enrolment, and 11 of these 138 (8.0%) tested positive. All previously rRT-PCR positive participants were IgA and IgG seropositive at the baseline time point of our study. Conversely, 16/126 (12.7%) and 5/126 (4.0%) previously rRT-PCR negative participants had tested positive for IgA or IgG, respectively. During the period between the initial (March 16-April 3) and baseline (April 15-April 29) testing (mean interval between tests: 26.9 days, range: 13-37 days), viral clearance, shown by a negative rRT-PCR at baseline, occurred in 10/11 (90.9%), while a single participant remained positive at baseline, 25 days after initial testing (see above).

Among all IgG positive participants (n=35), only one participant reported travelling within the previous 14 days and three reported having contacts outside the household (confinement measures were already in place since March 16, 2020). Seven participants were working on-site while 13 were working from home. Thirteen (37.1%) participants reported having contact with confirmed or suspected COVID-19 cases, most of the time (11/13, 84.5%), this person being household members (**Table 6**).

**Table 6:**
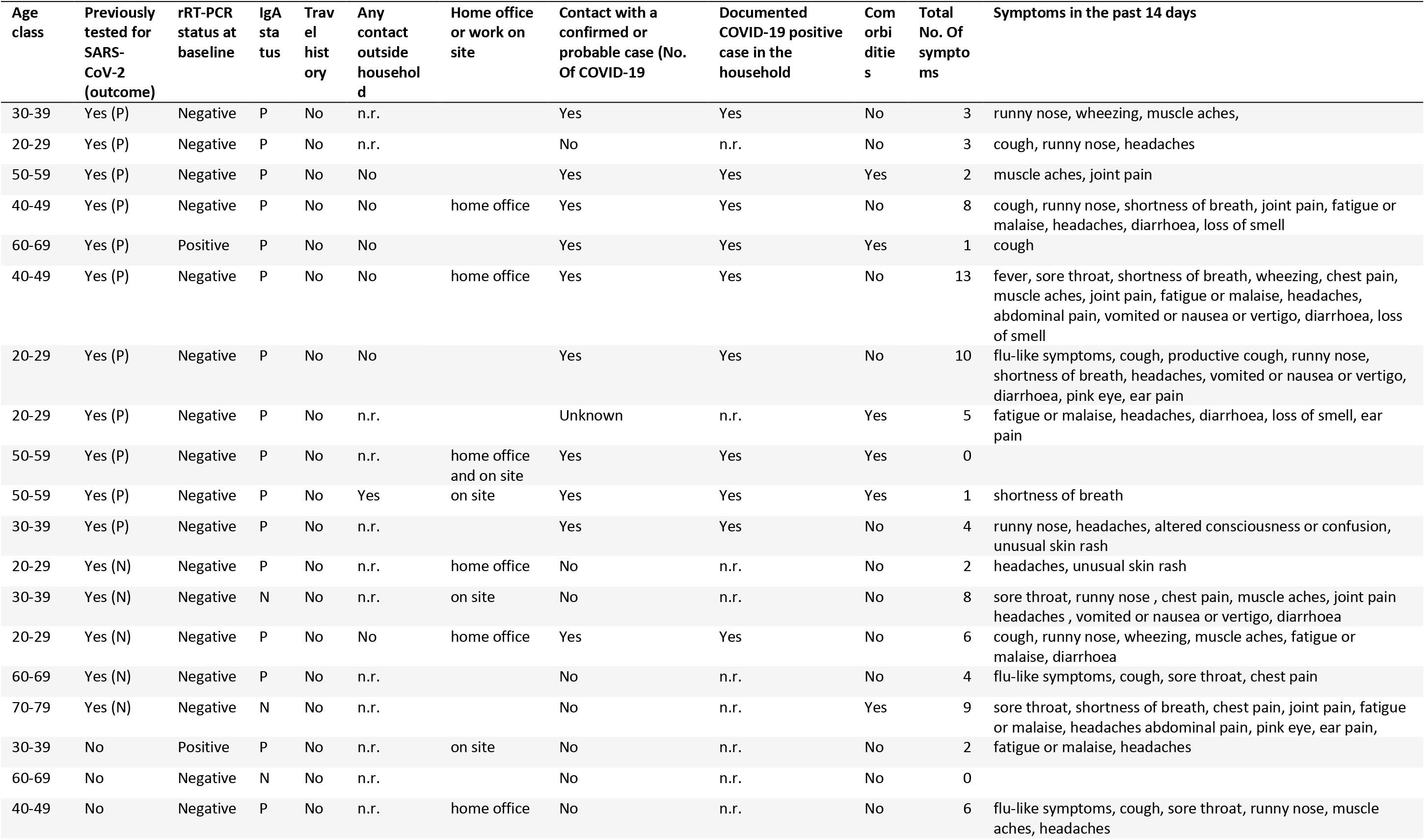
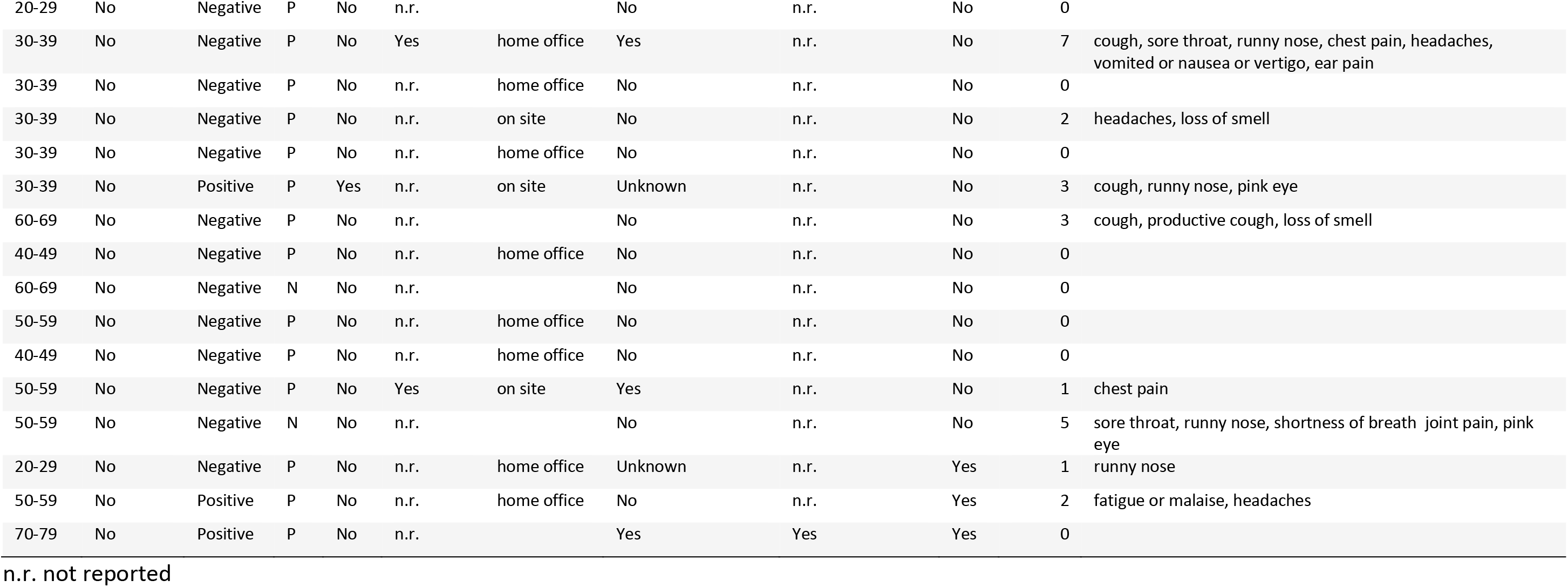
Overview of contact (any and with COVID-19 cases), travel history to regions with documented COVID-19 circulation, reported comorbidities and self-reported symptoms in the last 14 days in IgG seropositive participants (n=35)

A wide range of clinical symptoms were reported for the last 14 days prior to enrolment by IgG positive individuals. Symptoms ranged from none or very mild (≤2 symptoms in 19 participants) to more substantial symptoms, including shortness of breath, chest and abdominal pain. The number of symptoms reported tended to be higher in participants with a previous rRT-PCR positive diagnostic (range 0-13, average 4.5) than in participants with no or a negative rRT-PCR result (range 0-9, average 2.5; **Table 6**). The number of symptoms reported by participants with comorbidities (n=9; average 2.3) tended to be lower than in participants with no reported comorbidities (n=26; average 3.5).

### Prevalence evaluation

For the prevalence evaluation, two out of 1842 participants with rRT-PCR results were excluded because the gender was not defined. Participants aged > 79 years were also excluded (n=12) because representativity for the overall population was not reached for this age group. The target population for prevalence evaluation was therefore defined as people between age 18 years until age 79 years. The number of individuals with rRT-PCR and serology results available was 1830 and 1807, respectively with 4 participants negative in serology with no PCR results.

The sampling frame was used to weight the number of individuals in each cell defined by the three stratification variables. Weights were calculated by dividing each cell count from the target population by the one from the sample. It was therefore used in post-stratification.

The evaluations of prevalence used the weights to extrapolate to the target population and evaluate the 95% confidence intervals.

The time prevalence of SARS-CoV-2 RNA by rRT-PCR was 0.30% (95%CI=[0.03;0.56]) during the period of April 16 until May 5. This result translates into 1449 (95%CI=[145;2754]) people in the general population aged 18 years to 79 years.

Seroprevalence of IgA was 11.07% (95%CI=[9.54;12.60]) and that of IgG 2.09% (95%CI=[1.37;2.82]). The Infection rate was 2.06% (95%CI=[1.34, 2.77]) (**Table 7**).

**Table 7:** Prevalence estimates with weights calculated with gender, 10 years age categories and electoral district

| Positivity | Frequency | Estimate (%) | 95% CI |  | Design Effect |
| --- | --- | --- | --- | --- | --- |
|  |  |  | Lower Limit (%) | Upper Limit (%) |  |
| RT PCR | 5/1830 | 0.2973 | 0.0298 | 0.5649 | 1.1478 |
| IgA | 200/1807 | 11.073 | 9.5404 | 12.6057 | 1.1199 |
| IgG | 35/1807 | 2.0947 | 1.3683 | 2.8212 | 1.2082 |
| Infection rate (IR) | 35/1835 | 2.0586 | 1.3446 | 2.7726 | 1.2055 |

As a sensitivity analysis, weights were recalculated using Luxembourg’s regional cantons rather than electoral districts, a more granular geographic representation. An electoral district is composed of 2 to 5 cantons. Participants were randomly selected in the district, but as we also obtained the canton information, it was relevant to have an estimate of how the prevalence could be biased by potential clustering within districts.

Using the more granular canton approach, the time prevalence of SARS-CoV-2-rRT-PCR was 0.32% (95%CI=[0.02;0.63]) which was an estimate very close to that obtained at the district level. A similar observation could be made for the other estimated prevalence (**Supplementary Table 2**).

## Discussion

The present study provides first prevalence data on the spread of SARS-CoV-2 in the population of Luxembourg using a panel-based approach. The weighted prevalence of SARS-CoV2 carriers based on rRT-PCR reveals a low prevalence of 0.30%, which may be due to the confinement measures in place for more than 7 weeks, including social distancing for the overall population and self-isolation and quarantine. Our data suggest that between April 16 and May 5 there were 1,449 adults in Luxembourg that were oligo- or asymptomatic carriers of the SARS-CoV-2.

Similarly, low results from population screenings in Iceland were recently reported with 0.8% virus-positive individuals in an open-invitation screening and 0.6% in a random-population screening identified (Gudbjartsson et al., 2020). The relevance of assessing the contribution of asymptomatic and oligosymptomatic individuals to the dynamics of the pandemic is increasingly recognized and also motivated a study at the National Institute of Health (USA) using a similar population-based approach where participants fill questionnaires during a virtual clinical visit and are biosampled for assessing seropositivity (NCT04334954).

Of the five SARS-CoV-2 positive participants only one reported that he had travelled to a COVID-19 risk area. Two positive participants reported prior contact with a COVID-19 diseased household member. The source of infection for the other 2 SARS-CoV-2 positive participants is not clear, but most probably within Luxembourg, as travels were limited given the confinement measures in place.

For the presence of infected individuals within a household, we observed the highest percentage in households with 2 members and the lowest percentage in households with 5 or more individuals (**Suppl. Fig. 1**). This is in line with the observation in a recent German study that showed a relatively moderate increase of the secondary infection risk dependent on the household cluster size (Streeck et al., 2020).

Overall participants tested positive for an active infection of SARS-CoV-2 by rRT-PCR, were oligo-or asymptomatic and presented with fewer symptoms (mean of 1.6 symptoms out of 22; SD=1.14) than individuals with negative rRT-PCR results (mean of 2.3 symptoms out of 22; SD=2.43). This underscores the relevance of infected individuals that do not display typical COVID-19 signs and symptoms for the viral spread during the pandemic.

In line with previous reports, we see males overrepresented in the group of SARS-CoV-2 positive participants (4 males versus 1 female) (Gudbjartsson et al., 2020; Guan et al., 2020). As we targeted a population >18 years old, we cannot make any direct statement on the contribution of children to the viral spread.

All of the data was collected through online questionnaires. In highly connected countries such as Luxembourg, approximately 97% of the population accesses and regularly uses the internet (International Telecommunications Union, 2019). Previously, self-reported online questionnaires have been shown to be a reliable method of accessing the clinical evolution of disease (Davies 2016), and the lack of face-to-face interaction, flexibility to complete the questionnaires at the participants’ convenience, together with the perceived degree of anonymity inherent in such online questionnaires is thought to enhance data accuracy, reducing central coherence and social desirability biases at the cost of participants potentially misinterpreting the questions (Ong and Weis, 2000).

Retrospective assessment was limited to events and symptoms occurring in the previous two weeks to reduce recall bias. The rapidity with which such a self-reporting system that was accessible throughout the population was rolled out was crucially important in the pandemic situation. This rapidity means that reliable data can be provided promptly to researchers during the ongoing pandemic. Furthermore, in such an unpredictable, rapidly changing pandemic situation follow-up questionnaires can be modified as quickly as ethical approval can be obtained. The advantages were, however, counterbalanced by the possibility of survey fatigue with five survey waves in quick succession.

The geographical location of Luxembourg and the nature of the study required that several compromises were made. Luxembourg is highly multilingual and successfully recruiting a representative cohort required that study participants were offered the choice of questionnaires in French, German, English and Portuguese. The problems associated with translating questionnaires are well known (Pan and Fond 2014), and to circumvent this, we restricted the study to questionnaires that had previously been experimentally validated in all languages. As we aimed to detect asymptomatic SARS-CoV-2 carriers the ISARIC case report form was adapted to participants that had limited or no symptoms and the Luxembourgish sociodemographic situation.

There are pros and cons of using web panels for surveys. We used the services of a private company for immediate access to potential participants from all over the country and based on prior knowledge on age, gender and residency. The sample of participants to the current study was enrolled through the use of a non-probabilistic web panel (unknown probability to opt-in) of 18,000 panel members. Registration to the panel was constituted via invitation during telephone or face to face interviews as well as ad banners, media campaigns and homepage of the company. The sample was randomly drawn from the panel for each stratum defined by the crossing of the 3 stratification variables through a deterministic random bit generator (DRBG) within strata. Individuals living across the borders were excluded as we aimed at a sample of Luxembourgish residents.

The advantages of using such a panel was the simplification of the logistics to implement the study in a short time frame in order to capture the prevalence of the infection (**Figure 6**) while the decreasing slope of the epidemics has already started. Another aspect to consider is that no hospitalised patient was included in our sample as they were not reachable through the panel.

**Figure 6:**
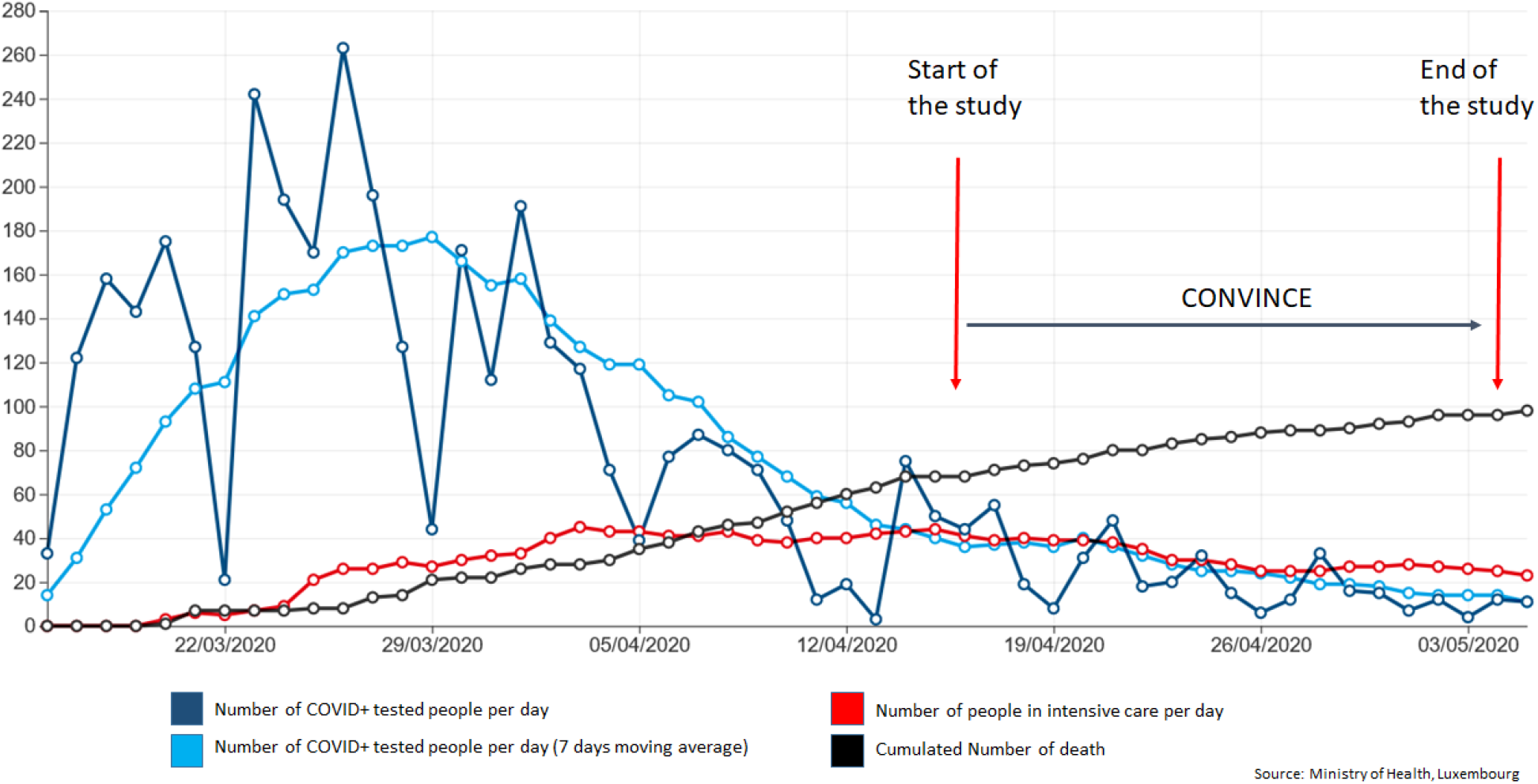
Course of the infection in Luxembourg until 5th May 2020 (Number of people COVID+ tested, in intensive care or death per day)

However, online surveys may be biased samples because the respondents are self-selected whereby individuals who have strong opinions, are overrepresented, and individuals that are indifferent or apathetic are less likely to opt-in or respond. Nevertheless, we tried to correct the selection bias and under representation by first stratifying the sampling on gender, age and residency at the sampling level and second, by post stratification at the analyses level.

Overall, our study captured 6 rRT-PCR positive participants with low viral RNA levels. These may be considered negative in other studies depending on the criteria applied. The combination of retesting by additional rRT-PCRs (**Figure 3**) as well as assessment of IgA and IgG serology, however, strongly suggest that 5 of these 6 study participants were true SARS-CoV-2 positive cases with residual low viral loads and seroconversion. This is also further evidenced by one of the participants who already tested positive 25 days prior to enrolment (participant 3, see **Table 4**). Duration of SARS-CoV-2 viral shedding appears to vary widely between patients. Detection of viral RNA for 2-3 weeks after symptom onset is frequent (Kujawski et al., 2020; Wölfel et al., 2020), even in seroconverted patients, and extended viral shedding for up to 83 days was reported (Li et al., 2020). One of the study participants (participant 5, **Table 4**) was not confirmed positive after two confirmation rRT-PCR assays and did not show seroconversion until now. This patient might have been recruited at the onset of viral shedding in contrast to the other cases and/or may not mount an immune response as sometimes observed in asymptomatic patients. Longitudinal follow-up sampling as implemented in the design of our study will allow us to clarify the status of this participant at a later stage.

The frequency of seasonal hCoV infections in the general population, including adults (Monto et al., 2020), coupled with possible concomitant infections by different hCoV (Heimdal et al., 2019) suggest a limited duration of protection for each hCoV and limited cross-protection between hCoV. Nevertheless, cross-reactivity in ELISA due to past exposure to other hCoV likely partially accounts for the observed lower specificity of the anti-SARS-CoV-2 IgA ELISA used in our study. Notably, cross-reactivity in anti-SARS-CoV-2 IgA and IgG ELISAs has been documented in patients with specific seroconversion against hCoV-OC43, while baseline sera were negative (Okba et al., 2020). In Germany, the follow up of a birth cohort revealed that 19/25 (76.0%) newborns had seroconverted by 21 months of age (Dijkman et al., 2012). While the frequency of each of the four seasonal hCoV oscillates between years and regions, hCoV-OC43 (betaCoV) is usually more frequent, in patients with acute respiratory disease, especially compared to hCoV-229E (alphaCoV) (Dijkman et al., 2012; Heimdal et al., 2019; Monto et al., 2020). Aside from a lower assay specificity, discrepancies between IgA and IgG responses may also arise from sequential immune response. IgA tends to appear earlier than IgG after symptom onset (Guo et al., 2020), and may explain why some participants have detectable levels of IgA but no IgG.

In this study, intermediate IgA and IgG antibody levels were assimilated to a positive serological response, as done previously (Streeck et al., 2020). Although this approach decreases specificity, the current study design allows for a careful monitoring of seroconversion trends and antibody levels over time during subsequent biosampling of study participants, which will allow us a better interpretation of evolving antibody responses. Additional immunoassays including alternative antigens or antigen fragments as well as neutralization assays will complement the panel of tests that will be performed at a later stage of the CON-VINCE study. These additional immunoprofiling data will provide a more refined assessment and evaluation of the ELISA assay results obtained at baseline level of our study.

### Outlook

The longitudinal design of our study with bi-weekly follow-ups will allow us to assess the dynamics of the pandemic in our study population along with the gradual easing of the protective measures taking place in Luxembourg as well as other European countries. This may inform about the impact of oligo- and asymptomatic carriers on the viral spread during the upcoming months. Moreover, we will be able to study immune responses via continuous serological testing and may provide information on the risk of re-infections, as knowledge about SARS-CoV-2 immunity is still limited.

## Data Availability

Due to ethical concerns, supporting data cannot be made openly available.

## Acknowledgments

We would like to give special thanks to all participants of the CON-VINCE study. Additionally, we are very grateful for the financial support by the Fonds National de la Recherche (FNR) and the André Losch Foundation, which enabled us to carry out the project. The funders had no role in the design and conduct of the study, nor in the decision to prepare and submit the manuscript for publication.

We would like to thank the Research Luxembourg COVID-19 Task Force (Frank Glod, Paul Wilmes, Lars Geffers, Jasmin Schulz, Henri-Cauchie, Ulf Nehrbass) and Rudi Balling for their overall support of the CON-VINCE study.

Furthermore, we would like to acknowledge the Communication team of the Research Luxembourg COVID-19 Task Force (Sabine Schmitz, Arnaud D’Agostini, Didier Gossens) for their excellent work and support during the implementation of CON-VINCE.

We would like to thank Philippe Lamesch for important and successful fundraising for research on COVID-19 in Luxembourg.

We thank the European Commission for the support of the European Virus Archive Global platform and the provision of SARS-CoV-2 material.

Furthermore, we acknowledge the joint effort of members of the CON-VINCE Study Group as listed below:

Tamir Abdelrahman, Geeta Acharya, Gloria Aguayo, Pinar Alper, Wim Ammerlaan, Ariane Assele Kama, Christelle Bahlawane, Katy Beaumont, Nadia Beaupain, Lucrèce Beckers, Camille Bellora, Guy Berchem, Fay Betsou, Luc Biever, Dirk Brenner, Eleftheria Charalambous, Emilie Charpentier, Estelle Coibion, Sylvie Coito, Manuel Counson, Brian De Witt, Antonella Di Pasquale, Palma Di Pinto, Olivia Domingues, Claire Dording, Jean-Luc Dourson, Bianca Dragomir, Thibault Ferrandon, Ana Festas Lopes, Manon Gantenbein, Piotr Gawron, Laura Georges, Soumyabrata Ghosh, Stéphane Gidenne, Georges Gilson, Enrico Glaab, Clarissa Gomes, Borja Gomez Ramos, Valentin Groues, Wei Gu, Gael Hamot, Anne-Marie Hanff, Linda Hansen, Maxime Hansen, Lisa Hefele, Ahmed Hemedan, Estelle Henry, Margaux Henry, Sascha Herzinger, Laetitia Huiart, Alexander Hundt, Gilles Iserentant, Anne Kaysen, Fédéric Klein, Tommy Klein, Stéphanie Kler, Rejko Krüger, Pauline Lambert, Sabine Lehmann, Anja Leist, João Manuel Loureiro, Andrew Lumley, Annika Lutz, François Massart, Patrick May, Monica Marchese, Sophie Mériaux, Maura Minelli, Joel Mossong, Friedrich Mühlschlegel, Maeva Munsch, Mareike Neumann, Beatrice Nicolai, Markus Ollert, Claire Pauly, Laure Pauly, Lukas Pavelka, Joëlle Penny-Fritz, Magali Perquin, Achilleas Pexaras, Marie France Pirard, Jean-Marc Plesseria, Laurent Prévotat, Guilherme Ramos Meyers, Kavita Rege, Lucie Remark, Antonio Rodriguez, Kirsten Rump, Estelle Sandt, Bruno Santos, Venkata P. Satagopam, Aurélie Sausy, Christiane Schmitt, Margaux Schmitt, Reinhard Schneider, Valerie Schröder, Serge Schumacher, Alexandra Schweicher, Jean-Yves Servais, Florian Simon, Chantal Snoeck, Kate Sokolowska, Lara Stute, Stéphane Tholl, Noua Toukourou, Johanna Trouet, Christophe Trefois, Nguyen Trung, Jonathan Turner, Michel Vaillant, Carlos Vega Moreno, Charlène Verschueren, Claus Vögele, Maharshi Vyas, Xinhui Wang, Femke Wauters, Bernard Weber, Tania Zamboni

## Additional Information

ClinicalTrials.gov number, NCT04379297

## Supplementary Figure

**Supplementary Figure 1:**
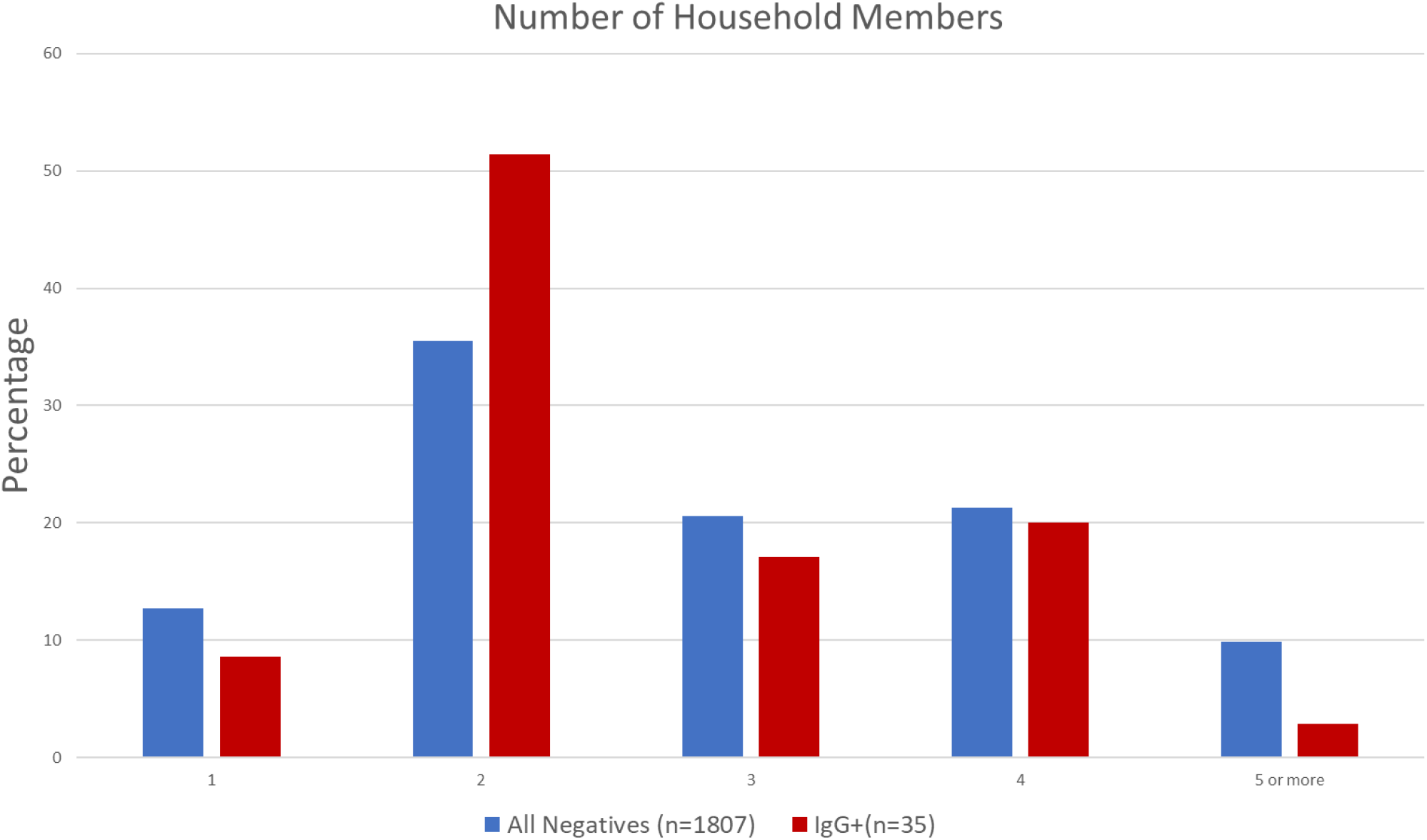
Number of household members (including the participant) for participants tested IgG positive compared to participants tested RT-PCR and IgG negative (All Negatives).

**Supplementary Table 1:** Biosample collection, processing and storage

| Biosample | Primary Container | Process | Derivative | Aliquoting Scheme | Storage Conditions |
| --- | --- | --- | --- | --- | --- |
| Nasal Swab<br>Oropharyngeal Swab | Dual Swab collection in 2ml Universal Transport Medium (UTM) | Swab/UTM aliquoting | Swab/UTM | 200 µl x 1<br>1.2 ml x 1 | -80°C |
|  |  | RNA extraction | RNA | 100 µl x 1 | -80°C |
| Blood | 10 ml CAT tube | Serum aliquoting | Serum | 400 µl x 6 | -80°C |
|  | 10 ml EDTA tube | Plasma aliquoting | Plasma EDTA | 400 µl x 6 | -80°C |
|  |  | Buffy coat aliquoting | Buffy Coat | 450 µl x 2 | -80°C |
| Stool | Zymo DNA/RNA Shield Collection Tube | Stool aliquoting | Stool | 500 µl x 4 | -80°C |
|  |  |  | Stool | 3.8 ml x 1 | -80°C |

**Supplementary Table 2:** Prevalence estimates with weights calculated with gender, 10 years age categories and canton

| Positivity | Frequency | Estimate (%) | 95% CI |  | Design Effect |
| --- | --- | --- | --- | --- | --- |
|  |  |  | Lower Limit (%) | Upper Limit (%) |  |
| RT PCR | 5/1830 | 0.3282 | 0.0208 | 0.6355 | 1.373 |
| IgA | 200/1807 | 10.7013 | 9.1691 | 12.2335 | 1.1533 |
| IgG | 35/1807 | 1.9705 | 1.2519 | 2.6891 | 1.255 |
| IR | 35/1835 | 1.9391 | 1.2316 | 2.6466 | 1.2551 |

## Supplementary Material 1

CON-VINCE CASE RECORD FORM Version 1.5 08 April 2020

Adapted from Sprint Sari Case Report Form by ISARIC. Used and made available by ISARIC under CC BY SA 4.0

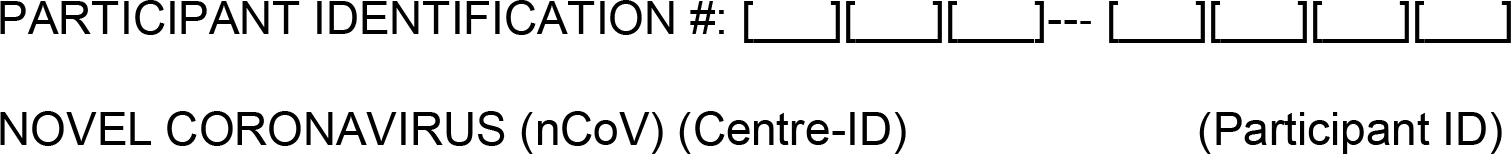

**CON-VINCE Study** – Electronic Case Record Form

(full extended part with defined minimal data set)

**EXCLUSION CRITERIA**

Infection by Coronavirus with severe disease course requiring a hospital admission prior to the inclusion to the study.

□ Yes

□ No

Presence of fever and respiratory distress/cough at the time of inclusion not attributable to other known chronic disease.

□ Yes

□ No

**Explanatory note for participant:** The virus called SARS-CoV-2 (further indicated as Coronavirus) is the cause of the current pandemic disease known as COVID-19. It is a rapidly spreading virus affecting mainly the respiratory tract having up to 80% an asymptomatic course (meaning without any symptoms) or very mild upper-respiratory disease with runny nose or pink eye. However, in a fraction of patients, the disease evolves to fever and cough with or without respiratory distress and in some cases, a hospitalisation with therapy by inhaled oxygen and further medical support is needed.

**(Minimal Data set) Coronavirus STATUS**

**Have you already been tested for Coronavirus?^*^**

□ Yes

□ No

*If yes*:

**When have you been tested for Coronavirus?**

[_D_][_D_]/[_M_][_M_]/[_2_][_0_][_Y_][_Y_]

*If yes*:

**What was the result of your Coronavirus test?**

□ Positive

□ Negative

□ Unknown

* *Replaced by the following question for the Follow-ups*:

**Since your visit to the lab for this study, have you been tested for Coronavirus? [outside the context of this study]**

**(Minimal Data set) EPIDEMIOLOGICAL FACTORS**

**In the last 14 days, have you travelled to a foreign country with documented cases of Coronavirus infection?^2^**

□ Yes

□ No

□ Unknown

*If Yes:*

**How many countries did you travel to?^2^**

Please specify: _________________________________________________________________

*If Yes:*

**Please document the following elements for each of your visits to a foreign country with documented cases of Coronavirus infection in the last 14 days.^2^**

*If you visited more than 5 countries, please refer to 5 most recent ones*.

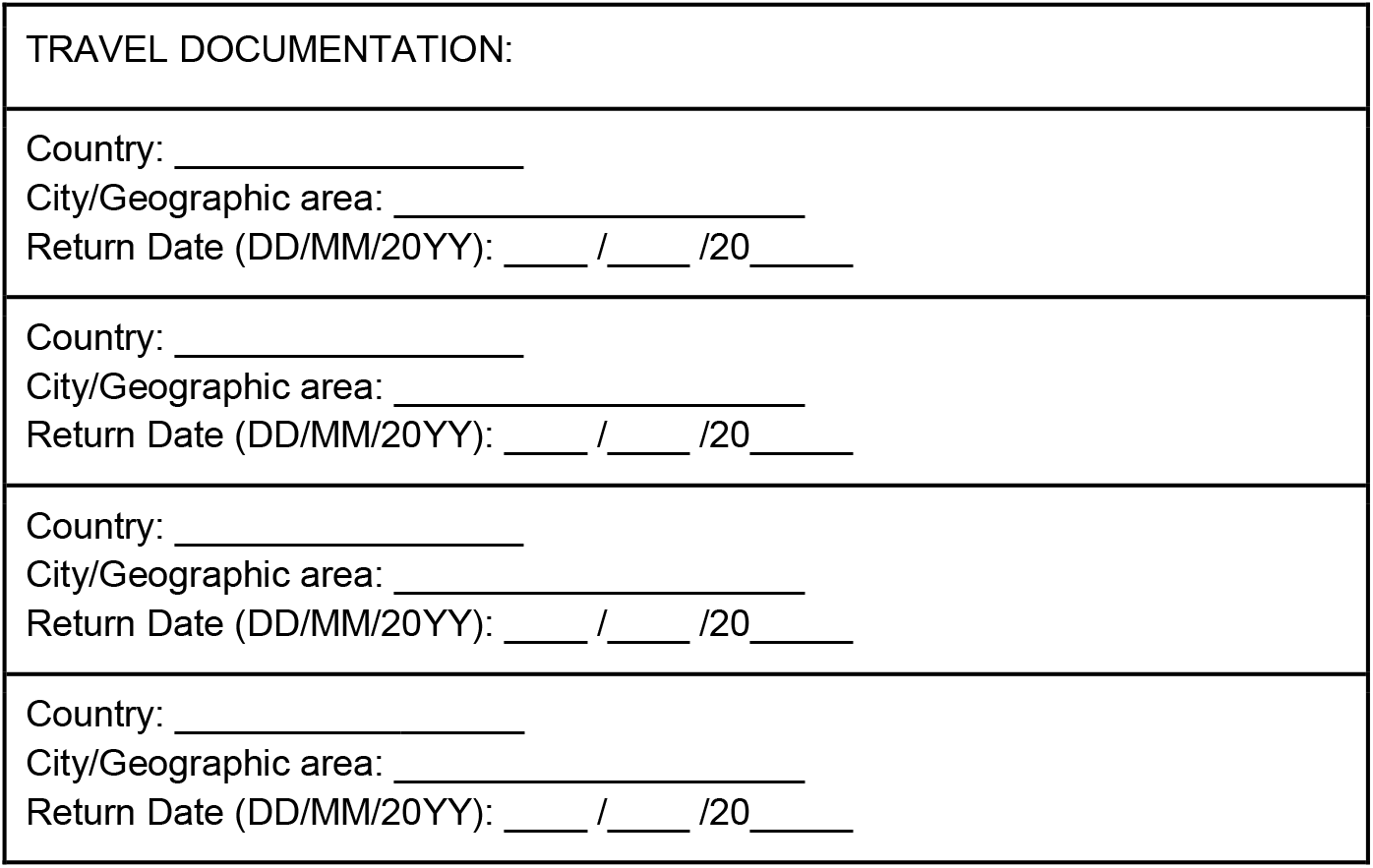

**In the last 14 days, have you been in close contact with a confirmed or probable case of Coronavirus infection, while that patient was either asymptomatic (having no symptoms such as fever/cough) or symptomatic (having symptoms such as fever, cough or respiratory distress)?**

□ Yes

□ No

□ Unknown

*Close contact is defined as:*

- *Health care associated exposure, including providing direct care for novel coronavirus patients, e.g. health care worker, working with health care workers infected with novel coronavirus, visiting patients or staying in the same close environment of a novel coronavirus patient, or direct exposure to body fluids or specimens including aerosols*.
- *Working together in close proximity or sharing the same classroom environment with a novel coronavirus patient*.
- *Traveling together with novel coronavirus patient in any kind of conveyance*.
- *Living in the same household as a novel coronavirus patient*

*If Yes:*

**Who was the person infected with Coronavirus (or believed to be infected) you have come to contact?**

□ Household member
□ Family member who does not live at home
□ Close neighbor
□ Friend
□ Co-worker
□ Cleaning staff, caregiver, household services, or similar
□ Coronavirus + patient (in the case of health personnel)
□ Professional client, customer
□ Public sector worker (e.g. cashier, bus driver, etc.)
□ Other contacts
□ Don’t know

**In the last 14 days, have you been present in a healthcare facility where Coronavirus infections are managed?**

□ Yes

□ No

□ Unknown

**In the last 14 days, have you been present in a laboratory handling suspected or confirmed Coronavirus samples?**

□ Yes

□ No

□ Unknown

**(Minimal Data set) DEMOGRAPHICS**

**Please insert in the data file the date of the interview**.

[_D_][_D_]/[_M_][_M_]/[_2_][_0_][_Y_][_Y_]

**What is your nationality?** *If you have dual or multiple nationalities, please don’t hesitate to indicate them at the next Question*.

Please specify (list of nationalities): _________________________________________________________________

**Do you have dual or multiple nationalities?**

□ Yes

□ No

*If yes:*

**Please indicate your second nationality**.

Please specify (list of nationalities):

**What is your country of origin?**

Please specify (list of countries): _________________________________________________________________

**Country of current residence = Luxembourg**

**In which municipality do you live?**

Please specify (list of municipalities in Luxembourg): _________________________________________________________________

**Please indicate your zip code**.

Please specify: L-_______

□ Unknown

**What is your gender at birth?**

□ Male
□ Female
□ Prefer not to say
□ Diverse

**Please indicate your current age in years**.

Years: ____________

**What is your current marital status?**

□ Single
□ Married
□ Registered partnership
□ Divorced
□ Widowed
□ Other status, please specify ____________

**How many children do you have?**

Please specify: _____________________ □ No children

**How many grandchildren do you have?**

Please specify: _____________________ □ No grandchildren

*Filter: only female respondents:*

**Are you pregnant at the moment?**

□ Yes

□ No

□ Unknown

*If Yes:*

**Please indicate at which gestational week you are pregnant**.

Week: ___________________

**Have you given birth in the last 6 months?**

□ Yes

□ No

□ No answer

*If Yes:*

**Please indicate the pregnancy outcome**.

□ Live birth

□ Still birth

□ No answer

*If Livebirth:*

**Please indicate the delivery date**.

[_D_][_D_]/[_M_][_M_]/[_2_][_0_][_Y_][_Y_]

*If Livebirth:*

**Has your baby been tested for Coronavirus?**

□ Yes

□ No

□ No answer

*If Yes:*

**What was the result of the Coronavirus-test?**

□ Positive

□ Negative

□ Unknown

**(Minimal Data set) EDUCATION / PROFESSIONAL BACKGROUND**

**What is your educational degree?**

*Please choose the highest degree achieved*.

□ No formal degree
□ Fundamental Education
□ Secondary Education - Classical system
□ Secondary Education - Technical system
□ University degree: Bachelor
□ University degree: Master or above
□ Other type of degree, please specify: ______________

**How many years of schooling have you successfully accomplished?**

*Please indicate in years [kindergarten not counted, and PhD counts as maximum 3 years]*

Years: ______________

**What is your current employment status^1^?**

□ Full-time employed
□ Part-time employed
□ Self-employed or working for own family business
□ Unemployed
□ In vocational training/retraining/education
□ Parental leave
□ In retirement or early retirement
□ Permanently sick or disabled
□ Looking after home or family
□ Other, please specify: ______________

*Filter: Full-time employed, part-time employed, self-employed or working for own family business, parental leave*

**What is your current professional activity?**

Please specify: __________________________

**In which field do you work?**

□ Essential services (e.g. police, firefighter)
□ Wholesale / retail trade
□ Manufacturing industry
□ Health and social services activities ^**^
□ Hospitality
□ Education
□ Public administration and defense
□ Construction
□ Transport and storage
□ Administrative activities and auxiliary services
□ Professional, scientific and technical activities
□ Agriculture, livestock, forestry and fishing
□ Information and communications
□ Domestic staff
□ Financial and insurance activities
□ Artistic, recreational and entertainment activities
□ Sanitation, waste management and decontamination activities^**^
□ Other services, please specify: __________________________

*Link ^**^ to this question:*

**Are you employed as a Healthcare Worker?**

□ Yes

□ No

□ Unknown

*If Yes:*

**More specifically, what is your profession?**

□ Nurse
□ Physician
□ Pharmacist
□ Physiotherapist
□ Occupational Therapist
□ Psychologist
□ Dietician
□ Secretary in healthcare
□ Cleaning staff in hospital/healthcare
□ Technician
□ Other profession, please specify: _______________________________

*Link ^**^ to this question:*

**Are you employed in a microbiology laboratory?**

□ Yes

□ No

*If Yes:*

**Have you been in contact with suspected COVID-19 positive samples?**

□ Yes

□ No

**(Extended part) HOME AND SOCIAL CONTACT / SOCIO-ECONOMIC STATUS**

**What is the type of your household?**

□ House
□ Apartment
□ Nursing home
□ Residence for the disabled
□ Jail
□ Hotel
□ Another type of shared residence (monastery, etc.)
□ Homeless
□ If none of the above, please specify: ________________

**How many people live in your household (including yourself)?**

□ 1
□ 2
□ 3
□ 4
□ 5 or more

**What is the age of household members (including yourself)? Please type in the number of the household members to the appropriate age categories**.

*If a category doesn’t apply, please indicate 0*.

Number of 0 - 4 years old [___]

Number of 5 - 9 years old [___]

Number of 10 - 14 years old [___]

Number of 15 - 19 years old [___]

Number of 20 - 29 years old [___]

Number of 30 - 39 years old [___]

Number of 40 - 49 years old [___]

Number of 50 - 59 years old [___]

Number of 60 - 69 years old [___]

Number of 70 - 79 years old [___]

Number of 80+ years [___]

**Is one of your household member coronavirus positive?^1^**

□ Yes □ No □ Unknown (not tested)

*If Yes:*

**What is the age of household members (yourself included) that have been tested coronavirus positive? Please type in the number of the household members to the appropriate age categories.^1^**

*If a category doesn’t apply, please indicate 0*.

Number of 0 - 4 years old [___]

Number of 5 - 9 years old [___]

Number of 10 - 14 years old [___]

Number of 15 - 19 years old [___]

Number of 20 - 29 years old [___]

Number of 30 - 39 years old [___]

Number of 40 - 49 years old [___]

Number of 50 - 59 years old [___]

Number of 60 - 69 years old [___]

Number of 70 - 79 years old [___]

Number of 80+ years [___]

**Do you own the home you live in?**

□ Yes

□ No

**What is your household gross annual income?**

□ 0 - 25 000 Euros
□ 25 000 - 50 000 Euros
□ 50 000 - 75 000 Euros
□ 75 000 - 100 000 Euros
□ 100 000 - 150 000 Euros
□ More than 150 000 Euros
□ No answer

**Are you in self-isolation / self-quarantine?^1^**

□ Yes

□ No

*If Yes:*

**Since when are you in self-isolation / self-quarantine?^1^**

*Please indicate the date*.

[_D_][_D_]/[_M_][_M_]/[_2_][_0_][_Y_][_Y_]

*Filter: Ask only if: Full-time employed, part-time employed, self-employed or working for own family business, parental leave, self-isolation/self-quarantine = No*

**Are you still traveling to work? ^1^**

□ Yes □ No

*If Yes:*

**With how many people do you currently share your office/workplace? ^1^**

□ I work alone at the office
□ 1-2
□ 3-5
□ 6 or more

*Filter: Ask only if: Full-time employed, part-time employed, self-employed or working for own family business, parental leave*

**Are you doing home-office? ^1^**

□ Yes □ No

*If Yes:*

**Since when are you doing home-office?^1^**

*Please indicate the date*.

[_D_][_D_]/[_M_][_M_]/[_2_][_0_][_Y_][_Y_] □ I’m always doing home office

**(Minimal Data Set) MAJOR LIFE EVENTS**

Turner, J. D., Ambrosio, C. D., Vögele, C., & Diewald, M. (2020). Twin Research in the Post-Genomic Era: Dissecting the Pathophysiological Effects of Adversity and the Social Environment. *International Journal of Molecular Sciences, 21*, 1-18.

**(Minimal Data set) COMORBIDITIES**

**Please indicate your current height (in cm)?**

Please specify: ______________________________

**Please indicate your current weight (in kg)?**

Please specify: ______________________________

**Are you affected by chronic cardiac disease** *(meaning chronic heart failure including inborn heart disease, but without including elevated blood pressure/hypertension*)*?*

□ Yes □ No □ Unknown

**Are you affected by a cardiovascular disease** *(e.g. heart infarction, Angina pectoris, placement of coronary stent)****?***

□ Yes, please specify: ______________________________ □ No □ Unknown

**Are you affected by hypertension** *(meaning: chronic elevated Blood pressure over 140/90 or treatment by antihypertensive medication)****?***

□ Yes □ No □ Unknown

*If Yes:*

**Do you take any antihypertensives?**

□ Yes, please specify the name of the antihypertensives: ______________________________ □ No

***Are you affected by chronic pulmonary disease*** *(chronic inflammatory lung disease- e.g. chronic obstructive pulmonary disease but without including asthma)****?***

□ Yes, please specify: ______________________________ □ No □ Unknown

**Are you affected by asthma** *official diagnosis by a physician)****?***

□ Yes □ No □ Unknown

**Do you have any type of Diabetes?**

□ Yes □ No □ Unknown

*If Yes:*

**Do you have any chronic diabetes-related complications** *(e.g. diabetic foot or chronic kidney disease due to the chronic diabetes)****?***

□ Yes □ No □ Unknown

**Do you suffer from chronic kidney disease** *(meaning long term decrease of kidney function or kidney failure)****?***

□ Yes □ No □ Unknown

*If Yes:*

**Are you on dialysis?**

□ Yes □ No □ Unknown

**Do you suffer from a rheumatologic disorder** *(meaning: inflammation that affects the connecting or supporting structures of the body* — *most commonly the joints, but also sometimes the tendons, ligaments, bones, and muscles)****?***

□ Yes, please specify: ______________________ □ No □ Unknown

**Are you affected by a moderate or severe liver disease** *(chronic liver dysfunction of any cause)****?***

□ Yes □ No □ Unknown

**Have you been diagnosed with dementia** (e.g. Alzheimer disease, vascular dementia, Lewy Body dementia)**?**

□ Yes, please specify: ______________________ □ No □ Unknown

**Have you had a stroke?**

□ Yes □ No □ Unknown

**Are you affected by a mild liver disease** *(chronic liver dysfunction of any cause diagnosed by a physician)****?***

□ Yes, please specify: ______________________ □ No □ Unknown

**Do you suffer from malnutrition?**

□ Yes □ No □ Unknown

**Are you affected by a chronic neurological disorder** *(e.g. multiple sclerosis, epilepsy, neuromuscular disorders)****?***

□ Yes, please specify: _________________________ □ No □ Unknown

**Are you affected by a neurodegenerative disease** *(e.g. Parkinson****’****s Disease)****?***

□ Yes, please specify: _________________________ □ No □ Unknown

**Have you been diagnosed with any form of cancer** (malignant neoplasm)?

□ Yes, please specify which type: _________________________ □ No □ Unknown

**Have you been diagnosed with Chronic hematologic disease** *(e.g. lymphoma, leukemia, multiple myeloma)?*

□ Yes, please specify: _________________________ □ No □ Unknown

**Have you been diagnosed positive for virus HIV** *(AIDS)?*

□ Yes □ No □ Unknown

**Are you affected by an autoimmune disease** *(e.g. autoimmune thyroiditis-Hashimoto disease, Sjögren syndrome, Lupus erythematosus)***?**

□ Yes, please specify: _________________________ □ No □ Unknown

**Have you undergone an organ transplantation?**

☐ Yes, please specify which organ: _______________ ☐ No ☐ Unknown

**Are you on any treatment suppressing your immune system** (e.g. immunosuppressive therapy for an autoimmune disease or chemotherapy)**?**

□ Yes, please specify: _________________________ □ No □ Unknown

**Do you suffer from any psychiatric disease** (e.g. depression, schizophrenia, bipolar disease)**?**

□ Yes, please specify: _________________________ □ No □ Unknown

**Are you affected by any other relevant disease not addressed before?**

□ Yes, please specify: _________________________ □ No

**Do you exercise <u>during</u> the Coronavirus pandemic?^1^**

□ Yes □ No

*If Yes:*

**How many hours <u>per week</u> do you exercise during the Coronavirus pandemic?^1^**

*Please indicate in hours. If a category doesn’t apply to you, please indicate 0*.

Hours inside: ___________________

Hours outside: ___________________

**Did you exercise before the measures implemented in response to the coronavirus pandemic?**

□ Yes □ No

*If Yes:*

**How many hours <u>per week</u> do you exercise before the Coronavirus pandemic?**

*Please indicate in hours. If a category doesn’t apply to you, please indicate 0*.

Hours inside: ___________________

Hours outside: ___________________

**Do you currently smoke?**

□ Yes
□ No, but I am a former smoker
□ No and I never smoked
□ No, but I live with someone who smokes

**(Minimal Data set) CURRENT MEDICATION**

**Do you take medication on a regular basis?**

□ Yes □ No

*If Yes:*

**How many different medications do you take on a regular basis?**

Please indicate a number: _________

**Please document the following elements for every medication you take on a regular basis**.

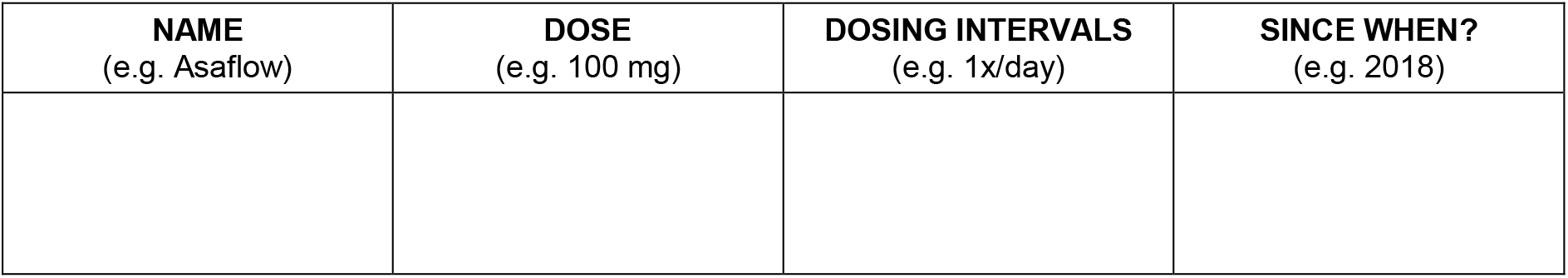

**(Minimal Data set) RESPIRATORY SYMPTOMS ONSET**

**Do you currently have flu-like symptoms (e.g. fever, cough, runny nose, pink eye)?**

□ Yes □ No □ Unknown

*If Yes:*

**When did the first/earliest flu-like symptom(s) occur?**

Please indicate the date.

[_D_][_D_]/[_M_][_M_]/[_2_][_0_][_Y_][_Y_]

**Do you take any anti-inflammatory medication or any medication to reduce fever or pain*****(e.g. Ibuprofen, paracetamol, aspirin or other)?***

□ Yes, please specify: _________________________ □ No □ Unknown

**Signs and symptoms (initial symptoms) (observed/reported by participant and associated with this episode of acute illness, meaning the occurrence of the symptoms in the recent 14 days)**

**In the last 14 days, have you…**

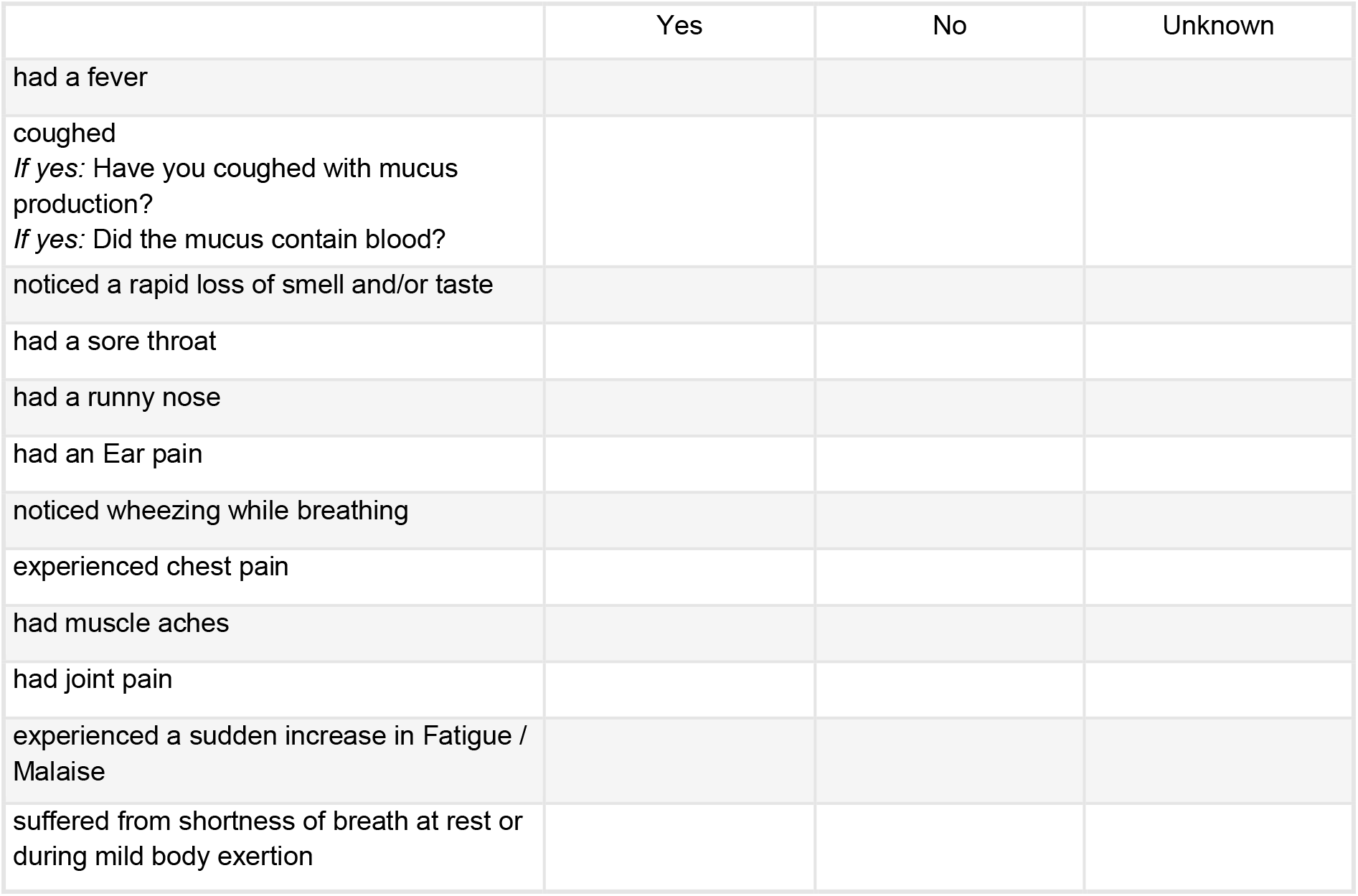

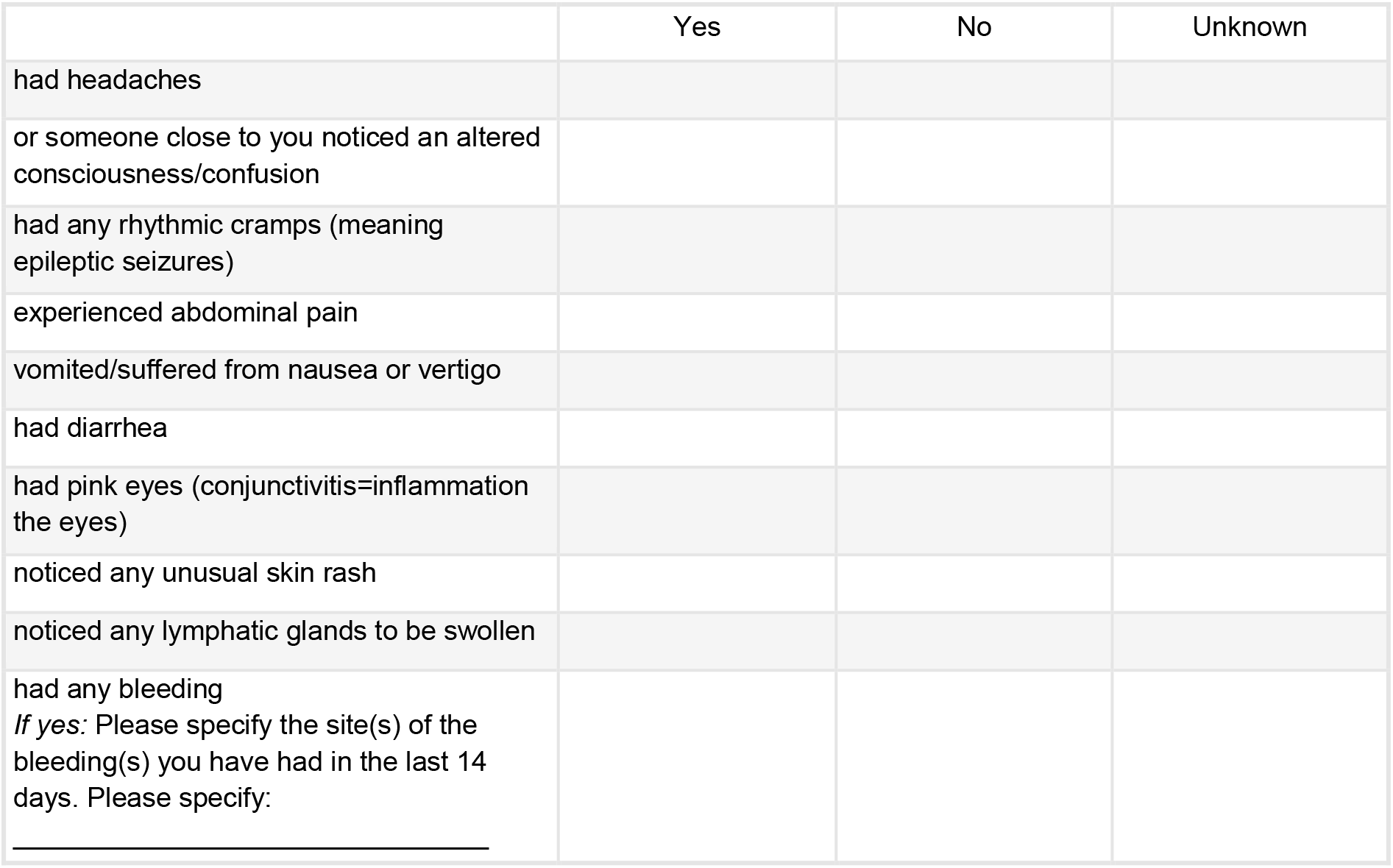

**Extended part) ENVIRONMENTAL CONDITIONS AT HOME WITH A DIAGNOSIS OF COVID-19**

*Filter: Asked only if COVID positive OR if living with a COVID positive household member^3^*

**Do you currently dispose of…?**

**Do you currently dispose of ...?**
|  | Yes | No | Unknown |
| --- | --- | --- | --- |
| a separate room for the patient |  |  |  |
| the necessary medication |  |  |  |
| food |  |  |  |
| masks for the patient |  |  |  |
| masks for the family members providing care |  |  |  |
| gloves |  |  |  |
| hand hygiene products |  |  |  |
| of a caregiver |  |  |  |
| access to medical advice and care |  |  |  |

**Is there currently a person at risk living at your home?**

*A person at risk is defined either by age (over 60) and/or being affected by coronary heart disease/ hypertension/ diabetes/ chronic pulmonary disease and/ or having compromised immune system by an immuno-suppressive therapy or due to an immuno-compromising disease*.

□ Yes □ No □ Unknown

**(Extended part) BEHAVIOR ANALYSIS DURING CORONAVIRUS - PANDEMIC / COMPLIANCE TO THE NATIONAL GUIDELINES**

**Are you familiar with the recommendations and restrictions issued by the government in Luxembourg?**

□ Yes □ No

**To what extent do you agree with the following statements?**

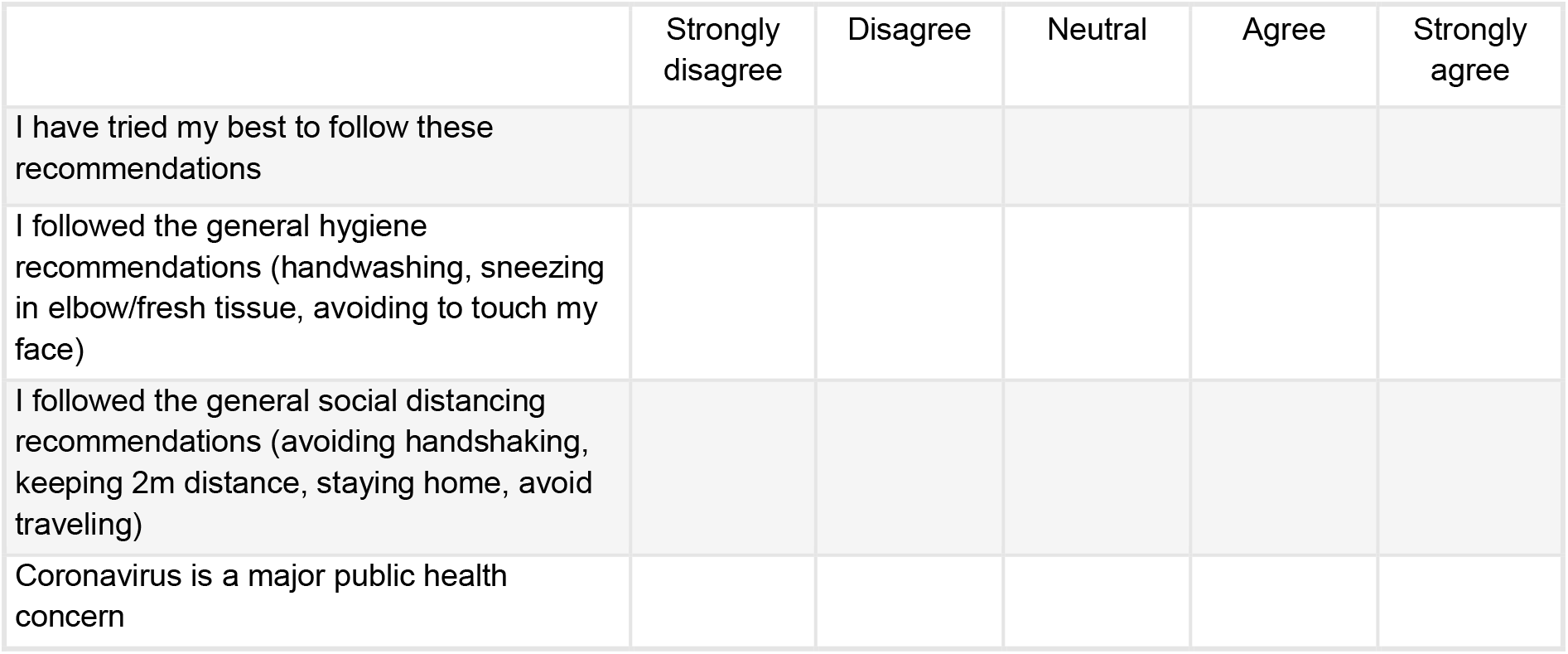

**How often did you leave the house <u>during the last week</u>**?

*If you didn’t leave the house during the last week, please indicate 0*.

Please indicate in days: ______________________ □ Unknown

**How often did you leave the house <u>during an average week before the coronavirus pandemic</u>?^2^**

*If you didn’t leave the house during the last week, please indicate 0*.

Please indicate in days: ______________________ □ Unknown

**Do you drink alcohol?**

□ Yes □ No

**How many units of alcohol did you drink <u>during the last week?</u>**

*Hereby a few examples of 1 unit: Large shots of spirit (35ml); Bottle of beer (330ml); Standard glass of wine (175ml)*

Please specify: ______________________ □ Unknown

**How many units of alcohol did you drink <u>during an average week (before the Coronavirus pandemic)</u>**?^2^

*Hereby a few examples of 1 unit: Large shots of spirit (35ml); Bottle of beer (330ml); Standard glass of wine (175ml)*

Please specify: ______________________ □ Unknown

**Over the last two weeks, how many hours <u>per day</u> have you spent…**

|  |  |
| --- | --- |
| surfing the internet? | _____hours/day |
| on social media platforms (e.g., Facebook, Instagram, Twitter)? | _____hours/day |
| watching news on television? | _____hours/day |
| speaking with friends or family (via telephone, video calls etc.)? | _____hours/day |
| on streaming platforms (e.g., Netflix, Youtube)? | _____hours/day |

**(Extended Part) PSYCHOLOGICAL QUESTIONNAIRES**

---

|  | N° of items<br>(+optional<br>filter items) | Baseline | Follow-up 1 | Follow-up<br>2 | Follow-up<br>3 | Follow-up<br>4 | Annual<br>Follow-up |
| --- | --- | --- | --- | --- | --- | --- | --- |
| **Inclusion / Exclusion**<br>**Criteria** | 2 | 2 min | - | - | - | - | 2 min |
| **Coronavirus Status** | 1 (+2) | <1 min | <1 min | <1 min | <1 min | <1 min | <1 min |
| **Epidemiological factors** | 4 (+ 4) | 2 min | 2 min<sup>2</sup> | 2 min<sup>2</sup> | 2 min<sup>2</sup> | 2 min<sup>2</sup> | 2 min |
| **Demographics** | 11 (+6) | 1 min | - | - | - | - | 1 min |
| **Education /**<br>**Professional**<br>**background** | 3 (+4) | 2 min | <1 min<sup>1</sup> | <1 min<sup>1</sup> | <1 min<sup>1</sup> | <1 min<sup>1</sup> | 2 min |
| **Home and Social**<br>**Contact / SES** | 6 (+4) | 2 min | 1 min<sup>1</sup> | 1 min<sup>1</sup> | 1 min<sup>1</sup> | 1 min<sup>1</sup> | 2 min |
| **Major life events** | 12 | - | - | - | - | 3 min | - |
| **Comorbidities** | 31 | 3 min | - | <1 min<sup>1</sup> | <1 min<sup>1</sup> | <1 min<sup>1</sup> | 3 min |
| **Current Medication** | 1 (+ number<br>medication) | 1 min | - | - | - | - | 1 min |
| **Respiratory Symptoms**<br>**onset** | 1 (+1) | <1 min | <1 min | <1 min | <1 min | <1 min | <1 min |
| **Signs and Symptoms**<br>**during the last 14 days** | 24 (+3) | 2 min | 2 min | 2 min | 2 min | 2 min | 2 min |
| **SUB-TOTAL MINIMAL DATA**<br>**SET** | **96 (+24)** | **16 min** | **6 min** | **6 min** | **6 min** | **9 min** | **16 min** |
| **Environmental**<br>**conditions at home** | 10 | 1 min<sup>3</sup> | <1 min<sup>3</sup> | <1 min<sup>3</sup> | <1 min<sup>3</sup> | <1 min<sup>3</sup> | <1 min<sup>3</sup> |
| **Behavioral analysis**<br>**during COVID-19**<br>**Pandemic / Compliance** | 11 (+3) | 3 min | 2 min<sup>2</sup> | 2 min<sup>2</sup> | 2 min<sup>2</sup> | 2 min<sup>2</sup> | 2 min<sup>2</sup> |
| **CES-D (Depression)** | 20 | 10 min | 10 min | 10 min | 10 min | 10 min | 10 min |
| **GAD-7 (Anxiety)** | 7 | 1 min | 1 min | 1 min | 1 min | 1 min | 1 min |
| **UCLA short version**<br>**(Loneliness)** | 3 | <1 min | <1 min | <1 min | <1 min | <1 min | <1 min |
| **PSS/4 (Perceived**<br>**Stress Scale)** | 4 | <1 min | <1 min | <1 min | <1 min | <1 min | <1 min |
| **BRS (Resilience)** | 6 | 1 min | - | - | - | - | 1 min |
| **BFI-10 (Personality)** | 10 | 1 min | - | - | - | - | 1 min |
| **Childhood Trauma**<br>**Questionnaire** | 28 | - | - | - | - | 3 min | - |
| **SUB-TOTAL EXTENDED**<br>**PART** | **99 (+3)** | **18 min** | **14 min** | **14 min** | **14 min** | **17 min** | **14 min** |
| **TOTAL** | **195 (+27)** | **34 min** | **20 min** | **20 min** | **20 min** | **26 min** | **32 min** |

x^1^: Only the items marked with an ^1^need to be repeated at the Follow ups

x^2^: The items marked with an ^2^need to be asked only at Baseline

x^3^: This questionnaire needs to be asked only once (only at the moment when positive diagnosis for participant and/or for one of the household member)

